# Framework to estimate the cost-effectiveness of the Genome Sequencing-based surveillance network: an integrated operational model-epidemiological model approach

**DOI:** 10.64898/2026.07.11.26351795

**Authors:** Mayank Jha, Konduri Naga Abhishek Reddy, Nimalan Arinaminpathy, Abha Mehndiratta, Javier Guzman, Sripad Devalkar, Sarang Deo

**Affiliations:** Indian School of Business; Imperial College of London; Center for Global Development

**Author notes:** Correspondence and requests for materials should be addressed to SD.

## Abstract

Understanding how genomic surveillance capacity translates into population health outcomes is critical for designing effective pandemic response systems, yet the interaction between operational design and epidemiological dynamics remains insufficiently characterized. We develop an integrated analytical framework that links a whole-genome sequencing (WGS)-based surveillance network with a two-variant epidemiological transmission model to evaluate how surveillance operations influence variant detection, intervention timing, and health outcomes.

The framework combines a modified susceptible–exposed–infectious–recovered–susceptible (SEIRS) model with a detailed operational representation of a centralized WGS surveillance network in India, incorporating sample collection, transport, batching, sequencing capacity, and reporting delays. We simulate 54 scenario combinations defined by three sequencing capacity levels, three sampling proportions, three variant emergence timings, and two variant profiles (high severity–high immune escape and low severity–low immune escape). Detection of a novel variant triggers a modeled intervention consisting of isolation of some diagnosed individuals, increased testing rates across disease states, and expanded access to hospitalization.

Across simulations, the time from variant emergence to intervention implementation ranged from 73 to 351 days, depending on operational and epidemiological conditions. Increasing sampling proportion reduced detection time only when sequencing capacity was sufficient; under constrained capacity, higher sampling increased congestion and delayed detection. Expanding capacity from low to nominal levels substantially reduced turnaround times, with diminishing returns at higher capacity. Earlier detection consistently improved intervention effectiveness, with deaths averted ranging from 0.06% to 14.49% across scenarios. The cost per life-year saved ranged from INR 9,137 to INR 326,714 across all configurations, remaining below one to three times India’s GDP per capita, consistent with established cost-effectiveness thresholds.

These results demonstrate that the performance of genomic surveillance systems is jointly determined by operational and epidemiological dynamics. Effective surveillance design, therefore, requires coordinated optimization of sampling strategies and sequencing capacity to enable timely intervention and maximize population health benefits.

## 1. Introduction

More than 775 million infections and 7 million deaths have been reported due to SARS-COV-2 as of May 2024 [1]. Genomic surveillance played an essential role in early outbreak detection, tracking pandemic progress, devising intervention measures, and tracking intervention effectiveness. Whole Genome Surveillance (WGS) has been used in epidemiological surveillance of pathogens and assists in clinical decision-making on infectious diseases [2].

Usage of WGS for surveillance has increased in capacity and efficiency across the globe during the COVID-19 pandemic [3] with more than 15.5 million sequences uploaded on GISAID till September 8, 2023[4] and more than 18 variants and subvariants identified till May 2024 [5]. However, there is an apparent disparity in the percentage of samples sequenced. High-income countries like the United States of America and the United Kingdom sequenced 4.69% and 12.09% of their positive cases. In comparison, lower-middle-income countries (LMIC) like India and Egypt have sequenced merely 0.6% and 0.97% of their total positive cases[4]. Furthermore, the latter have experienced much longer Collection to Submission Time (CST) than the former [6].

One of the possible reasons for this disparity is the requirement for substantial investment in infrastructure and human capital [7]. The health outcomes from public health interventions like early variant detection or new therapeutic or vaccine development are based on the information derived from genomic surveillance programs [8]. Hence, the costs are apparent, but the benefits of WGS-based surveillance are much harder to quantify.

WGS-surveillance-based Infection Prevention program is less costly and more cost-effective than the Standard of Care in detecting hospital outbreaks [9]. WGS-based surveillance of healthcare-associated bacterial pathogens is overall cost-saving and more life-saving than current practices [10]. Genomic surveillance is a cost-effective intervention in controlling the spread of hospital outbreaks of methicillin-resistant Staphylococcus aureus (MRSA) [11].

In this study, we focus on a national-level pathogen genomic surveillance network and the impact of operational parameters like capacity or sampling proportion of the genomic surveillance network on novel variant detection time and total costs incurred. As shown in **Figure 1** we formulate a framework to evaluate the cost-effectiveness of a Genome-sequencing surveillance program by integrating surveillance network operations and the transmission dynamics of the pathogen. The daily count of positive samples from the epidemiological model is an input for the operational model to estimate the time of variant detection and intervention implementation.

**Figure 1:**
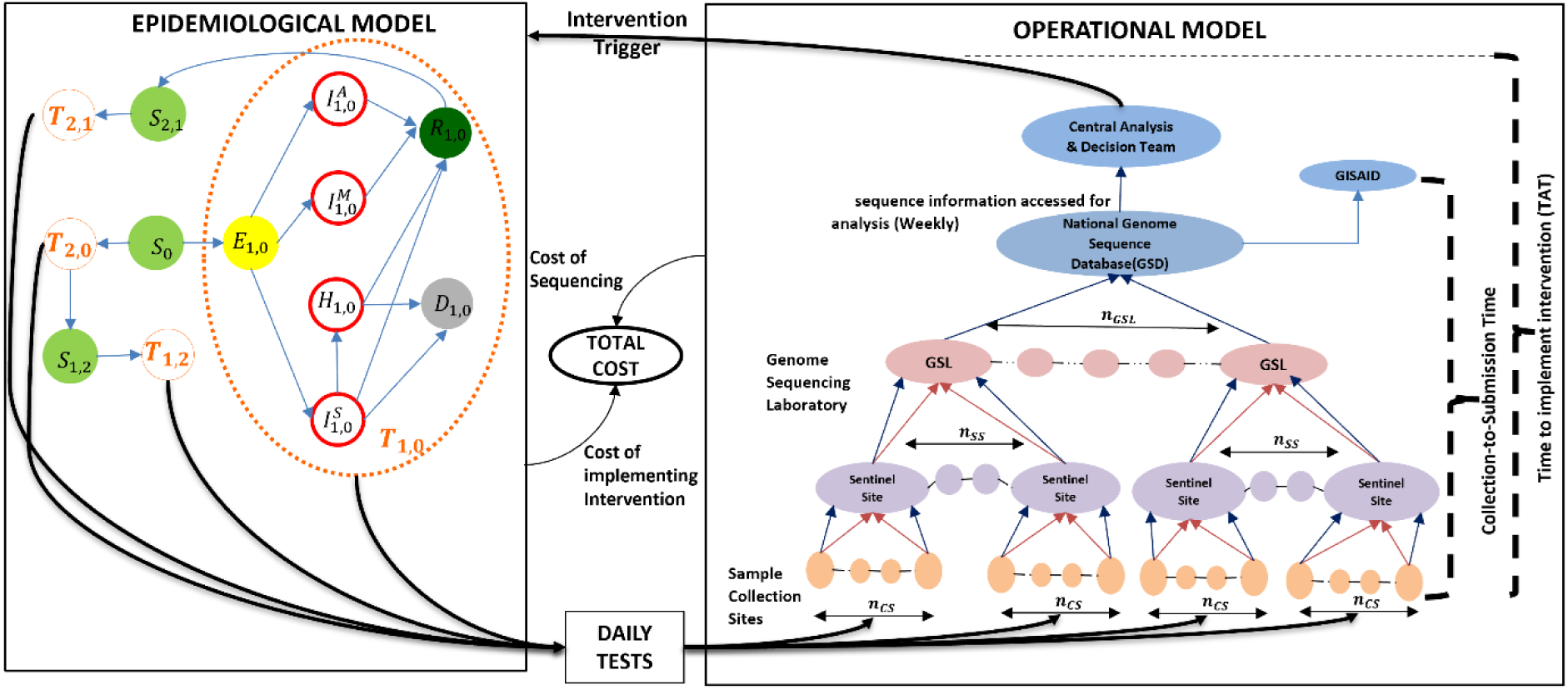
Integrated Epidemiological Model with two-variant transmission and Genome Sequencing Surveillance operational model. Note: This figure shows the integrated model of genome sequencing operations and two-variant transmission dynamics. *S*_0_ is the initial susceptible population, and’*S*_*x*,*y*_‘are susceptible to variant *x* after recovering from variant y infection where y=0 means no prior infection. ‘*T*_*x*,*y*_’ is the population progression between epidemic compartments exposed (*E*_*x*,*y*_), Infectious-Asymptomatic 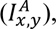 Infectious-Moderately Ill 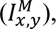 Infectious-Severely Ill 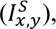 Hospitalized (*H*_*x*,*y*_), Recovered (*R*_*x*,*y*_) and Deceased (*D*_*x*,*y*_). The daily count of samples submitted by the Infectious population is input into the operational model. In the operational model, the red arrow denotes the physical transfer of samples, and the blue arrow represents information transfer, such as sample information. *n*_*CS*_ represents the number of samples per sentinel site, *n*_*SS*_ represents the number of samples per Genome Sequencing Lab (GSL) and ‘*n*_*GSL*_’represents the total number of Genome Sequencing Labs in the network. We model a centralized network where all samples are sent to a single Lab (*n*_*GSL*_ = 1) having the number of machines equivalent to the total number of machines in the decentralized network.

We modelled a centralized operational network of the Genome Sequencing Lab (GSL) operational network for WGS-based SARS-COV-2 surveillance based on visits to sequencing labs and interviews with subject experts. The operational model had the following components: a) sample collection and transportation between collection sites, sentinel sites, and sequencing labs, b) sequencing of samples at the Lab, and c) information flows between various nodes in the WGS ecosystem. Public health institutions implement interventions upon detection and subsequent Variant of Concern (VoC) analysis. The time of intervention implementation is an input to the epidemiological model. The cost-effectiveness of genome sequencing surveillance operations is calculated from the total cost of sequencing and intervention and the life-years saved due to intervention.

Our analysis explores the relationship between sequencing capacity, sampling proportion of RT-PCR positive samples, variant detection time and variant characteristics. Our results show the effect of increasing capacity on variant detection, incremental costs incurred, and effectiveness of sequencing networks in terms of life-years saved after intervention implementation.

## 2. Integrated Epidemiological - Genomic Surveillance Network Model

### Epidemiological Model

We developed a two-variant modified SEIRS (Susceptible-Exposed -Infectious-Recovered-Susceptible) compartment model [12] with multiple transmission pathways (*T*_*x*,*y*_) to account for the co-circulation of two variants in a closed population and the possibility of reinfection across variants. As shown in **Figure 1**, we segregated the Infectious compartment into Infectious-Asymptomatic 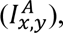 Infectious-Moderately Ill 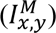 and Infectious- Severely Ill 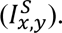 We also added a Hospitalized (*H*_*x*,*y*_) and Deceased (*D*_*x*,*y*_) compartment to capture the number of hospital admissions and deaths. We further augmented each disease state based on the patients’ state with regards to RT-PCR testing (test taken but waiting for results, received test results) and the test result (positive or negative) (see **Supplementary Figure 1**).

We assumed a closed-system transmission of pathogens with homogeneous inter-mixing of individuals, no births, and no deaths except due to infection. Individuals exposed to the pathogen become infectious after the latency period, either becoming asymptomatic, moderately ill, or severely ill. We assumed recovery for all asymptomatic and moderately ill individuals. We modelled hospitalization of severely ill individuals based on their hospital access, and those lacking access have a higher mortality rate than those with hospital access. We assume that infection from a variant gives lasting immunity against the same variant, and the individuals lose immunity and become partially susceptible to another variant upon recovery. The daily number of infectious individuals of each variant taking RT-PCR tests is an input to the operational model. Complete description of compartmental epidemiological model along and disease transmission equations of the model are in **Supplementary Section 1.1 – Supplementary Section 1.2**.

### Genomic Surveillance Operational Model

As shown in **Figure 1** under a decentralized operational network, the GSL are identical in terms of their operational characteristics (e.g., number of sentinel and collection sites associated with a lab, sequencing capacity, and frequency of sample transportation). Under a Centralized operational network, the number of sentinel sites and sequencing capacity associated with a centralized GSL is an amalgamation of all decentralized labs and sentinel sites. Hence, a centralized GSL has genome sequencing machines (GSMs) equivalent to the sum of all machines present in an analogous decentralized network. In this paper, we have modelled a centralized GSL with one lab where samples arrive from all sentinel sites.

In this model, individuals give their samples for RT-PCR testing at the sample collection sites at various public health facilities. We assumed an equal sample collection rate for RT-PCR testing across all collection sites and that 8% of all positively tested samples enter the WGS network [13]. At the end of each day, associated collection sites send samples to sentinel sites, and the sentinel site makes the test results available one day later. We modelled the uncertainty in collecting samples from individuals infected with the novel variant, considering that a surveillance network-associated collection site might not collect the novel variant sample (see **Figure 1**).

We modelled two types of sentinel sites: diagnostic laboratories and hospitals. Each diagnostic laboratory has six collection sites associated with it. A hospital acts as both a collection site and a sentinel site. Sentinel sites send a fraction of SARS-CoV-2 positive samples with a CT value less than 25 to GSL. We assumed each sentinel site sends samples weekly to centralized GSL We modelled the uncertainty in the likelihood of eligible samples of the novel variant being sent from a sentinel site to the GSL to capture the operational guideline’s impact on the timeline for novel variant sample arrival at a Sequencing Laboratory (see **Figure 1**).

We modelled the different processes within the sequencing lab to predict the timeframe between a sample’s arrival at the Sequencing Laboratories and the availability of the sequencing results (see **Supplementary Table 12 and Supplementary Table 13**). Based on information gathered during our interviews and field visits to one of the GSL, we noted that some of the activities in the sequencing lab are carried out in batches, i.e., multiple samples are processed together to save costs. To capture the impact of batch processes on the time taken, we modelled the time taken for enough samples to become available to form the required batch size and estimate the impact on total time due to batching.

Sentinel sites submit patient, laboratory, clinical, and travel information to a Genome Sequence Database (GSD). GSL conducts sequencing, analyses data, and uploads results to GSD. A central advisory team reviews GSD data for analysis and recommends public health interventions as needed. We assumed the central analysis team receives reports weekly, modelled as an average delay of 3.5 days from completing the sequencing to sharing the results for analysis. We also assumed an additional 15-day delay in implementing public health intervention. In our model, a central facility validates the genome sequences before uploading them to GISAID after sequencing. The Collection-to-Submission Time (CST) for each sequenced sample in the operational model includes time taken for data validation, the in-lab process at the sequencing lab, and an additional 4.5 days for submission.

### Model Calibration

In the Epidemiological model, Variant 1 transmission parameters corresponding to effective transmission rate, proportion of severely ill symptomatic infections, mortality rate in hospital, and recovery rate of hospitalized and non-hospitalized severely ill patients were calibrated to data for the diagnosed number of infections between February 2021 and December 2021 in the OWID dataset [14] (see **Supplementary Figure 2**) and the total estimated excess deaths [15]. Other Variant 1 transmission parameters are obtained from the literature as listed in **Table 1**.

**Table 1:**
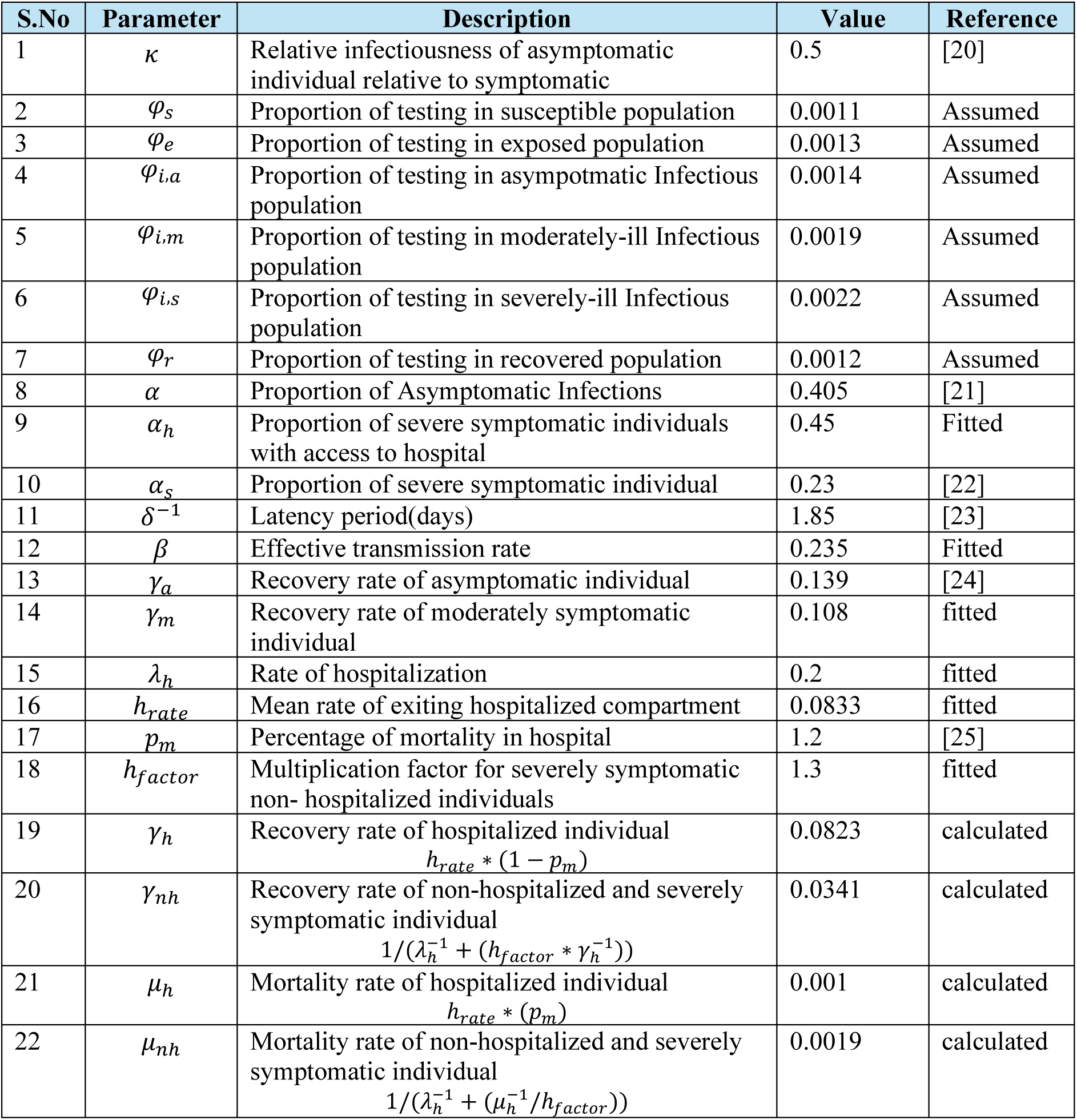
Transmission parameter values of Variant 1.

We calibrated sequencing capacity in the operational model to match the average CST of samples sent for sequencing each month to the reported month-wise CST estimate in [16] and the proportion of samples sequenced each week in the reported data.

The Epidemiological Model provides daily diagnosed COVID-19 case estimates from Feb 2021 to Dec 2021. Within each 7-day shipment cycle, the number of samples for sequencing is the product of the percentage of sequenced COVID cases per epidemiological week (EW) from Feb 2021 onwards [16] and the samples eligible for sequencing per week as obtained in the epidemiological model.

We used the GSAID data [16,17] of (i) the percentage of sequenced COVID-19 cases per epidemiological week (EW) between March 2020 and February 2022, (ii) the average collection to submission time (CST) of samples collected monthly, and the diagnosed cases from the transmission model to calibrate the operational model and estimate the sequencing capacity in India. The Mean Absolute Percentage Error (MAPE) estimates on the CST was approximately 11%, marking that the model estimates are reasonably closer to the reported data (see **Supplementary Figure 3**).

We estimate the batch size in the sequencing laboratories using the estimated sequencing capacity and the number of GSLs [18] between February 2021 and December 2021. The sequencing capacity was estimated to increase, indicating the expansion of the sequencing network with multiple labs with increased sequencing capacity in the population (see **Supplementary Figure 4**).

We estimated that by March 2022, the sequencing capacity per day in the sequencing network would be 6412. This capacity indicates that each of the 28 GSLs established in the sequencing network by March 2022 would have operated with one sequencer with a batch size 384. We assume that every Lab still operates with the same configuration, i.e., every 57 GSLs [18] operates with one sequencer with a batch size 384. In the epidemiological model, we estimated that a single variant transmission of Variant 1 results in 341489 samples/day at the peak of the epidemic cycle.

With 57 GSLs [18] in India as of July 2022, baseline sampling proportion of 5% of total positive samples [19] and assuming that Lab operates with same configuration, we defined Low Sequencing Framework (LSF) as a centralized network of 30 GSMs with utilization factor of 249%, Nominal Sequencing Framework (NSF) as a centralized network of 60 GSMs with utilization factor of 124% and High Sequencing Framework (HSF) as a centralized network of 90 GSMs with utilization factor 83%.

### Uncertainties and Scenarios

We simulated operational scenarios in the centralized network by changing the parameters of the sampling proportion of positive samples for sequencing (5%,10% and 30%) and in-lab operations, such as sequencing capacity (30 GSMs, 60GSMs and 90 GSMs), in a surveillance program. In the epidemiological model, we defined Variant Emergence Time (VET) as the number of days it takes for the novel variant (Variant 2) to emerge since the beginning of the simulation. We analyzed each operational scenario of the sequencing framework and sampling proportion across six epidemiological scenarios. These scenarios included two types of novel variant characteristics: high severity with high immune escape potential (HH) and low severity with low immune escape potential (LL). For each variant characteristic, we considered three VETs, representing different stages in the epidemic lifecycle of the original variant: early VET (25th day), near-peak VET (70th day), and late VET (160th day).

We addressed the uncertainty associated with pathogen transmission in the epidemiological model and novel variant detection in the operational model by employing an uncertainty analysis framework.

The transmission parameters for Variant 2 are derived by scaling the transmission parameters of Variant 1, using the multiplication factors contingent upon the defined epidemiological scenario. For the HH variant, we assumed an increase of 20% in transmissibility, mortality, and the proportion of severe infections and a decrease of 20% in recovery rates and the proportion of asymptomatic infections. For the LL variant, we assumed a decrease of 20% in transmissibility, mortality, and severe infection proportions and an increase of 20% in recovery rates and asymptomatic cases. We use Latin Hypercube Sampling (LHS) to draw multiplication factors from a normal distribution, with assumed means and coefficient of variation guiding the sampling process, as listed in **Table 2**. We assumed the coefficient of variation of the increase or decrease in recovery rate equal to the coefficient of variation of the increase and decrease in the proportion of severely ill patients. Each epidemiological scenario undergoes 30 iterations to capture a wide range of Variant 2 transmission parameters, ensuring the inclusion of the uncertainty inherent in the transmission dynamics.

**Table 2:**
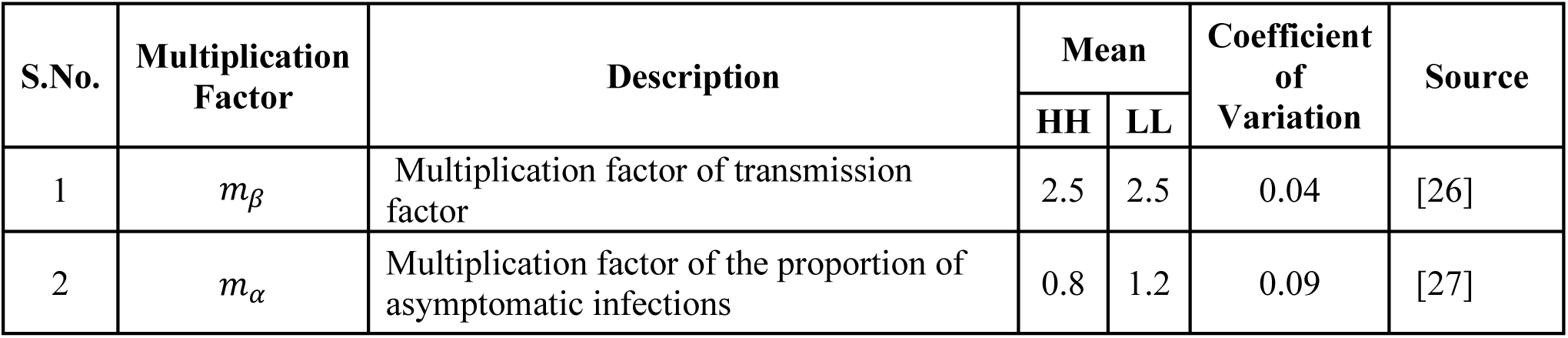
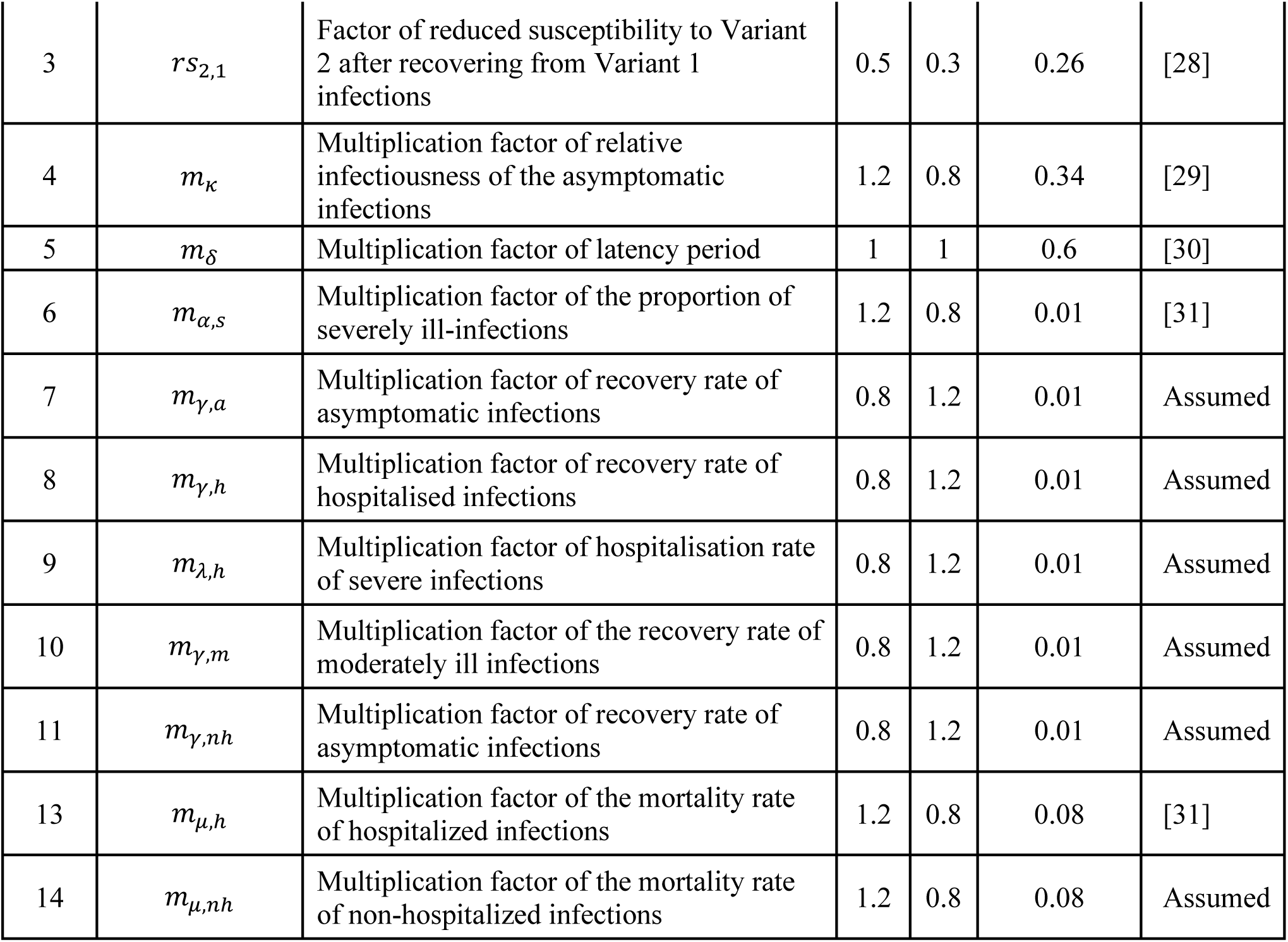
Multiplication factor for Variant 2 transmission parameters.

| S.No. | Multiplication Factor | Description | Mean |  | Coefficient of Variation | Source |
| --- | --- | --- | --- | --- | --- | --- |
|  |  |  | HH | LL |  |  |
| 1 | $m_\beta$ | Multiplication factor of transmission factor | 2.5 | 2.5 | 0.04 | [26] |
| 2 | $m_\alpha$ | Multiplication factor of the proportion of asymptomatic infections | 0.8 | 1.2 | 0.09 | [27] |
| 3 | $rs_{2,1}$ | Factor of reduced susceptibility to Variant 2 after recovering from Variant 1 infections | 0.5 | 0.3 | 0.26 | [28] |
| 4 | $m_{\kappa}$ | Multiplication factor of relative infectiousness of the asymptomatic infections | 1.2 | 0.8 | 0.34 | [29] |
| 5 | $m_{\delta}$ | Multiplication factor of latency period | 1 | 1 | 0.6 | [30] |
| 6 | $m_{\alpha,s}$ | Multiplication factor of the proportion of severely ill-infections | 1.2 | 0.8 | 0.01 | [31] |
| 7 | $m_{\gamma,a}$ | Multiplication factor of recovery rate of asymptomatic infections | 0.8 | 1.2 | 0.01 | Assumed |
| 8 | $m_{\gamma,h}$ | Multiplication factor of recovery rate of hospitalised infections | 0.8 | 1.2 | 0.01 | Assumed |
| 9 | $m_{\lambda,h}$ | Multiplication factor of hospitalisation rate of severe infections | 0.8 | 1.2 | 0.01 | Assumed |
| 10 | $m_{\gamma,m}$ | Multiplication factor of the recovery rate of moderately ill infections | 0.8 | 1.2 | 0.01 | Assumed |
| 11 | $m_{\gamma,nh}$ | Multiplication factor of recovery rate of asymptomatic infections | 0.8 | 1.2 | 0.01 | Assumed |
| 13 | $m_{\mu,h}$ | Multiplication factor of the mortality rate of hospitalized infections | 1.2 | 0.8 | 0.08 | [31] |
| 14 | $m_{\mu,nh}$ | Multiplication factor of the mortality rate of non-hospitalized infections | 1.2 | 0.8 | 0.08 | Assumed |

To account for the uncertainties in the sample collection and in-lab processes, we simulate 100 runs of the model and estimate the average turnaround time (from emergence to implementation of intervention) for each epidemiological iteration and surveillance network design combination. Detailed sample flow equations are present in **Supplementary Section 2.1**.

**Figure 2** shows the average time to implement intervention since variant emergence (TAT) with increasing sampling proportion for different VETs and variant characteristics at different Genome Sequencing Machines (GSMs). The time taken to identify a novel variant depends on combinations of operational policies such as the proportion of samples sent for sequencing, frequency of sending the samples for sequencing, operational batch size in the sequencing lab and the design of the network and characteristics of the novel variant.

**Figure 2:**
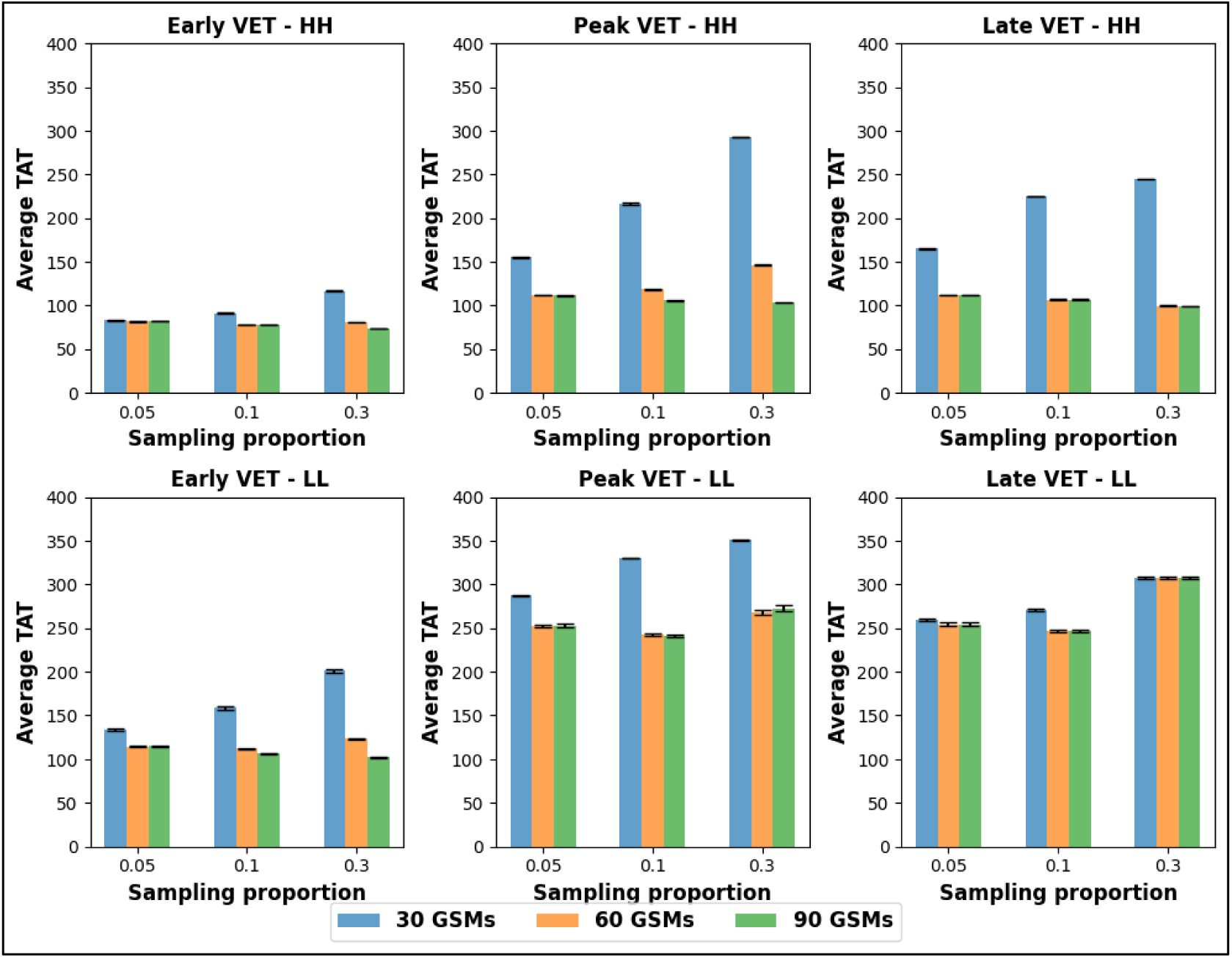
Average Turnaround Time (TAT) to implement an intervention. Note: This figure shows the average time taken to implement intervention (Average TAT) under a centralized genome sequencing network with 30, 60 and 90 Genome Sequencing Machines (GSMs) for epidemiological scenarios of early, peak and late Variant Emergence Time (VET) at sampling percentage of 5%,10% and 30%. This first row represents the Average TAT for the Novel variant of High Severity-High Immune escape potential (HH variant), and the second row represents the Average TAT for the Novel variant of the Low Severity-Low Immune escape potential (LL variant). Each subplot represents the Average TAT for a variant at a VET (early, peak or late) for operational configurations of sampling proportion and GSMs.

### Variant detection for different Variant characteristics

Variants are detected considerably quicker at early VET (VET = 25^th^ day) than at higher VETs (VET = 70^th^ day and VET = 160^th^ day). At VET =25^th^ day, TAT ranges from 73 to 201 days across variants. At peak and late variant emergence (VET = 70^th^ day and VET 160^th^ day), TAT ranges from 103 days to 351 days and 98 days to 308 days, respectively, across variants. Early VET variants are detected quicker than near peak, and late VET variants as fewer Variant 1 samples at the early transmission stage increase the probability of the first Variant 2 sample entering the sequencing network and facing lower wait time in the sequencing lab.

At peak and late variant emergence (VET=70^th^ day and VET=160^th^ day), the TAT of LL variant detection is considerably higher than the HH variant because of the lesser availability of samples in the network during the initial days of Variant 2 transmission. A variant with higher immune escape will infect a larger population, and consequently, more positive samples enter the sequencing network than a variant with lower immune escape potential. Similarly, a variant with higher severity will lead to more symptomatic infections and hospitalizations. Therefore, more positive samples are available in the sequencing network than a variant with lower severity.

### Variant detection due to changing sampling rate

In the Low Sequencing Framework (30 GSMs), an increase in sampling proportion leads to higher TAT ranging from an increase of 4.18% to 39.16% on increasing the sampling rate from 5% to 10% and an increase of 6.35% to 35.22% on increasing sampling rate from 10% to 30%.

In the Nominal and High Sequencing Framework, increasing the sampling rate from 5% to 10% TAT reduces 2.34% to 7.41% (except an increase of 5.95% in case of peak VET-HH variant in NSF). On increasing the sampling rate from 10% to 30% in NSF and HSF, the change in TAT ranges from a reduction in TAT of 3.88% (early VET-LL variant at HSF) to an increase in TAT of 23.56% (peak VET-HH variant at NSF).

Increased sampling proportions lead to faster arrival of novel variants at GSL. Depending on the congestion in the GSL, the queue time will then determine the variant detection time.

Sending more samples in a low sequencing framework increases the queue time for samples, leading to delayed variant detection. Nominal and high sequencing frameworks have enough capacity to process samples arriving at higher sampling rates, leading to TAT reduction.

Samples of the HH variant are available for sequencing much earlier than the LL variant. Increasing the sampling proportion in a peak VET-HH variant scenario leads to accumulating Variant 1 in the queue before the first HH variant sample can be sequenced. This increases TAT at a higher sampling rate in the Nominal Sequencing Framework.

At late VET, the effect of congestion in front of the first HH variant sample is less due to the shorter duration of co-circulation. As we increase the sampling rate beyond 5% at late VET, HH samples enter GSL quicker, and TAT decreases despite the accumulation of Variant 1 samples in GSL. This also indicates that increasing the sampling proportion beyond 30% may further decrease TAT.

### Variant detection at different Utilization Factor

The reduction in TAT at NSF (60 GSMs) in comparison to LSF (30 GSMs) ranges from 59.26% ( late VET-HH variant at 30 sampling rate) to 0.17% (late VET-LL variant at 30% sampling rate). Change in TAT ranges from a reduction of 29.6% (peak VET-HH variant at 30% sampling rate) to an increase of 1.64% (peak VET-LL variant at 30% sampling rate) on increasing capacity from NSF to HSF.

We observe that the impact of increasing capacity is more significant when more samples are sent for sequencing. The advantage of sending more samples is that the sample of Variant 2 will reach a GSL faster. By increasing capacity, we can observe the reduced wait time at GSL as more resources are available to process the samples waiting in the queue. However, the deployment of more resources leads to higher costs. When sending fewer samples, increased capacity may not be cost-effective as the baseline capacity is sufficiently large, so the waiting time inside GSL is low. Moreover, the GSL wait for samples to be collected to form a batch size. Hence, we also observe that increasing capacity and sampling proportion is considerably more effective when Variant 2 emerges near the peak (VET=70 days) of the Variant 1 transmission cycle, i.e., when the Variant 1 samples are much more than Variant 2 samples during the initial days of Variant 2 transmission.

The effect of increasing capacity decreases when we increase capacity from NSF to HSF, indicating that the capacity at 60 GSMs is adequate to detect new variants in most epidemiological scenarios. In a few epidemiological scenarios where the average TAT at HSF is higher than NSF, we find that the TATs at capacities lie within each other’s confidence interval.

## 3. Intervention upon variant detection

Detection of a novel variant in the operational network and its subsequent analysis triggers a public health intervention in the epidemiological model. We model public health intervention consisting of (i) isolation of 50% of diagnosed individuals, (ii) increased rate of testing amongst asymptomatic, moderately ill and severely ill individuals (explained in Section 2) by 20%, 50% and 100%, respectively, assuming that individuals in different disease states will be inclined to take COVID-19 tests differently, and (iii) increasing the proportion of the population having access to hospitals from 45% to 55%. The choice of intervention is based on interventions implemented in India during the SARS-COV-2 pandemic since 2020 [32]. These simulated intervention measures can be modified in the integrated framework depending on the stakeholders’ risk assessment of the detected variant.

As shown in **Figure 3**, we simulated the implementation of the intervention in every epidemiological scenario, with the time of intervention implementation ranging from the 50^th^ day to the 1000^th^ day since variant emergence time. Intervention effectiveness is calculated as the percentage of deaths averted due to intervention implementation. The percentage of deaths averted in an epidemiological scenario decreases with increasing time taken to implement the intervention. From **Figure 3**, we observe that the impact of the same intervention decreases at lower severity and immune escape potential of the novel variant. In every epidemiological scenario, the curve is flat for a period after which the percentage of deaths averted reduces drastically. Except for the early VET-HH variant scenario, in every epidemiological scenario, the duration for which there is no change in the percentage of deaths averted increases as severity and immune escape potential decrease.

**Figure 3:**
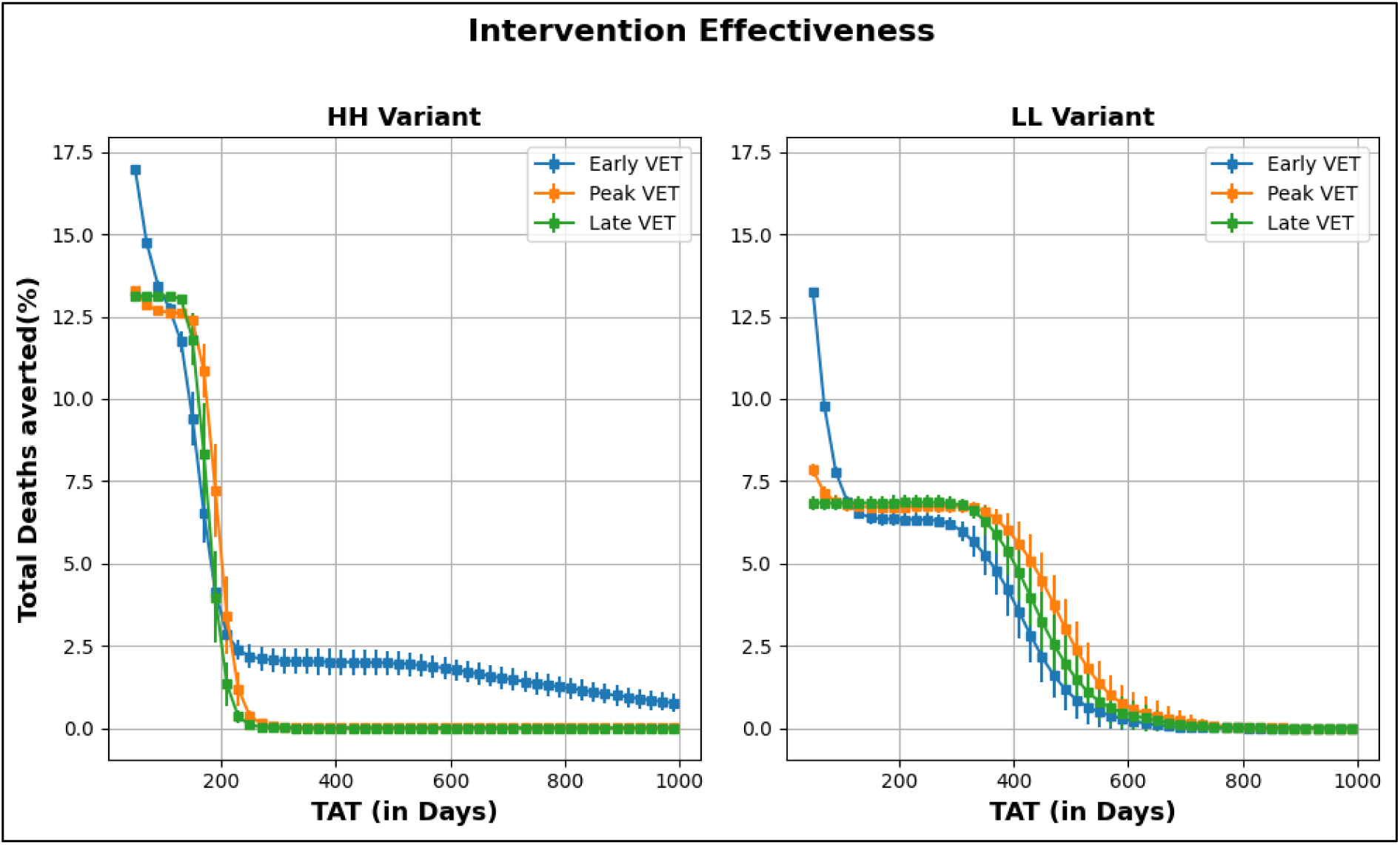
Intervention Effectiveness at Different Times of Intervention Implementation. Note: This figure represents the percentage of deaths averted due to intervention at different intervention implementation time (days). The time taken to implement intervention is the number of days since Variant emergence (VET). The left panel represents the percentage of deaths averted for High severity-High immune escape potential (HH variant), and the right panel represents the percentage of deaths averted for Low severity-Low immune escape potential (LL variant).

In the early VET-HH variant scenario, the intervention effectiveness drops faster than in other scenarios due to the quickest rise in cases. However, the percentage of deaths averted remains constant for a long duration before falling to zero. Though when Variant 2 emerges early, early interventions save more lives, in some iterations of this scenario, there is a second rise of infections after 600 days, which results in a positive percentage of deaths, which is averted even at a very delayed intervention time.

Intervention effectiveness in terms of the percentage of deaths averted with the same intervention is higher for the HH variant than the LL variant due to higher mortality in the HH variant in the absence of intervention. For a given VET, infections spread much quicker for a variant with higher immune escape potential and higher severity (HH). In any epidemiological scenario, the impact of intervention reduces once the infections rise exponentially. Hence, we see no decrease in the percentage of deaths averted for a longer duration in the case of the LL variant compared to the HH variant. For a given variant, an exponential increase in infections occurs much faster at early VET (VET = 25^th^ day) compared to VET = 70^th^ day and VET = 160^th^ day. Hence, intervention effectiveness decreases much earlier at VET =25^th^ day compared to peak and late VET(VET = 70th and VET = 160^th^ day) for respective variants.

The percentage of deaths averted with the same intervention is higher for the HH variant than the LL variant due to higher mortality in the HH variant in the absence of intervention. For a given VET, infections spread much quicker for a variant with higher immune escape potential and higher severity. In any epidemiological scenario, the impact of intervention reduces once the infections rise exponentially. Hence, we see no decrease in the percentage of deaths averted for a longer duration in the case of the LL variant compared to the HH variant. For a given variant characteristic, an exponential increase in daily infections occurs much faster at VET = 25th day compared to VET = 70^th^ day and VET = 160^th^ day. Hence, we observe that intervention effectiveness decreases much earlier at VET =25^th^ day compared to VET =70^th^ day and VET = 160^th^ day for respective variants.

## 4. Cost-effectiveness of Surveillance Network

The cost-effectiveness of an operational configuration should be interpreted as the effectiveness of an operational configuration associated with an intervention because the life years saved and the total costs incurred will vary with the intervention implemented.

We analyzed the performance of a genome sequencing operational network in terms of the total cost incurred in sequencing and implementing the intervention and the life-years saved after the intervention implementation. As described in Section 2, uncertainties in the operational network and the epidemiological model lead to different variant detection times and intervention implementation times at different iterations in any operational scenario. This results in different intervention effectiveness and the total cost incurred for an operational scenario at each iteration. The cost per life-year saved of an operational network in an epidemiological scenario is calculated as the mean of the ratio of the net cost of sequencing and intervention to the number of life years at each iteration. While we run each iteration of pathogen transmission for 2500 days with and without intervention, we calculate the end of pathogen transmission as the day when the number of hospital patients becomes zero. The total duration of pathogen transmission varies depending on the presence or absence of intervention. To facilitate a consistent comparison in each iteration of epidemiological model uncertainty, we calculate the cost of RT-PCR testing up to the maximum duration observed between the end time with and without intervention.

We estimated the average number of life-years saved on averting one death outside the model to be 19.98 years (see **Supplementary section 1.3**) based on the age-wise comparison of COVID deaths [33], the projected age-wise distribution of the population in 2021 from [34] and the expected life of an individual in an age-group from [35].

The net cost of public health intervention is estimated as the sum of the cost of quarantining, increased hospitalization access, and increased testing proportion of the infectious population. We obtained the per-day per-patient cost of monitored home isolation through personal interviews. We also estimated the cost of facility-based isolation and increased hospitalization by prorating the cost of care in Kenya [36] to India using critical-care costs in India [37]. A detailed calculation of the different costs is presented in **Supplementary section 1.4**. All costs of public health intervention were obtained in INR at the exchange rate of 1US $ = 82.67 INR as of June 5, 2023.

We also estimated the shipment cost (packing materials and transportation), personnel, reagents, equipment, and RT-PCR retest. We used Blue Dart courier service charges to calculate shipment costs in batches. We estimated personnel costs at GSL using information from the interviews and visits to GSL through activity-based costing. We used the cost of Illumina reagent kits [38] as reagent cost and collected each equipment item’s list and purchase cost from the interviews. Detailed calculations of the costs of each component of the operational network and the cost-per-sample estimates at different rates of sample arrival are in **Supplementary Section 2.2**.

The mean infection averted ranges from 0.01% (peak VET-HH variant in LSF at 30% sampling rate) to 0.52% (late VET-LL variant in LSF at 30% sampling rate) across the simulated epidemiological and operational scenarios. The percentage of deaths averted ranges from 0.06% (peak VET-HH variant in LSF at 30% sampling rate) to 14.49% (early VET-HH variant in HSF at 30% sampling rate) (see **Supplementary Figure 5 and Supplementary Figure 6**).

At a 5% sampling rate, the cost per life-year saved ranges from INR 9168.02 (late VET-LL variant in LSF) – 11108.85 (late VET-HH variant in LSF) across Low to High Sequencing Frameworks. The cost per life-year saved ranges from INR 9158.79 (late VET-LL variant in LSF)- INR 84531.71 (late VET-HH variant in LSF) and INR 9137.25(late VET-LL variant in LSF) – INR 326714.11(peak VET-HH variant in LSF) at 10% and 30% sampling rates, respectively, across all sequencing frameworks and epidemiological scenarios. As per WHO standards of cost-effective interventions, valuing a life year at one to three times GDP per capita [39], sequencing and subsequent intervention are highly effective.

As explained in **Section 3**, the number of monitored-home isolations and hospitalisations due to intervention are more significant in the HH variant scenario, resulting in higher costs and greater life-years saved compared to the LL variant (see **Figure 4**).

**Figure 4:**
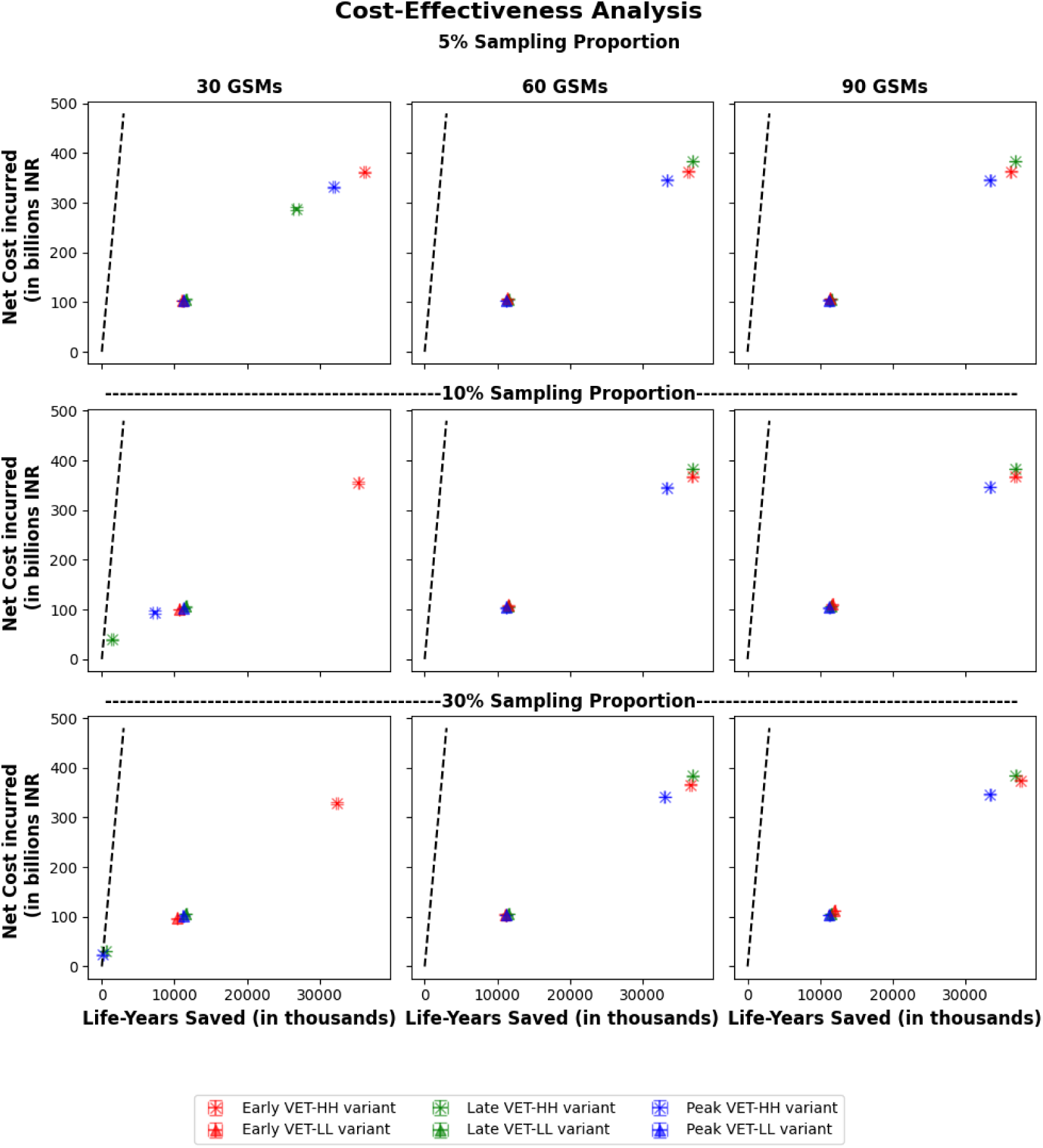
Cost-Effectiveness of operational network configuration. Note: This figure represents the cost-effectiveness of an operational configuration at a given number of Genome Sequencing Machines (GSMs) and the sampling proportion of positive samples for genome sequencing for different epidemiological scenarios. The x-axis is the number of life-years saved (in millions) due to intervention. The y-axis is the total cost of sequencing and implementing the intervention. The first sub-plot column represents costs and life-years saved at 30 GSMs, the second column at 60 GSMs and the third column at 90 GSMs. For a given GSM, the first row of subplots represents cost and life-years saved at a 5% sampling proportion, the second at a 10% sampling proportion, and the third at a 30% sampling proportion. Red markers in each plot represent High severity-high immune escape potential (HH variant), and green markers represent Low severity-low immune escape potential (LL variant).

For any epidemiological scenario, any significant change in total cost and number of life years saved happens only if a change in operational parameters leads to a significant delay in intervention and the intervention effectiveness has considerably reduced. We observed in **Section 3** that the number of deaths averted remains constant for a particular time duration and decreases rapidly later in each epidemiological scenario.

For the peak VET-HH variant, the total cost and number of life years saved are significantly lesser in LSF at a 30% sampling rate and increase if the sampling rate is reduced to 5% or 10% (see **Figure 4**). A similar trend is observed in the early VET–LL variant, where a higher sampling rate (30%) at LSF delays intervention implementation (see **Figure 2**) and reduces life-years saved and costs incurred.

Increasing the number of machines from 30 GSMs to 60 GSMs results in increased total costs and life years saved in the late VET-HH variant. When the novel variant wave is expected much later than the Variant 1 wave (late VET scenario), increasing capacity from NSF to HSF (60 GSMs to 90 GSMs) is ineffective, as there is no significant change in TAT.

## 5. Discussion

Whole genome sequencing, along with public health intervention, is a cost-effective measure over a range of epidemiological scenarios and operational configurations.

Increasing the sampling rate is cost-effective when the pathogen mutates frequently, as observed across all variants. This is because more samples are sent for sequencing, reducing the time taken to form batch sizes in the sequencing labs.

The lack of publicly available data regarding sequencing labs limits the operational model developed in this study. In reality, sequencing networks across the population will exhibit deviations from the model used. Centralized GSL, in reality, does not have aggregated resources but can collect and sequence a larger pool of samples. Further, we assumed that each GSM has a sequencer operating with a batch size of 384 to match the model calibration results. However, some GSMs will also operate with a batch size of 96. It is also possible that the in-lab operations in a decentralized network, which is assumed to be similar across all GSLs in the network, will vary. Also, we have assumed that the reagents used in the sequencing labs are supplied by ILLUMINA, which may not always be true, as various labs in a decentralized sequencing network may have contracts with other suppliers with lower unit costs than ILLUMINA.

The lack of more precise and granular data and information about sequencing labs prevented us from developing individual models that incorporated operational characteristics of the sequencing network for each decentralized GSL. While such granular models would have provided more precise estimates, they would not alter the directionality of critical findings or the implications for decision-making.

While a rise in cases may lead to some interventions or changes in individual behaviour without identifying a new variant, we have assumed that no intervention is implemented until the WGS-based surveillance network identifies a new variant. Our epidemiological model is limited by the assumption of homogeneous intermixing of individuals and no age stratification in susceptible compartments. However, while calculating the number of life years saved, we accounted for the age-wise distribution of deaths in India [33].

Since isolation-center and ICU costs are prorated from the costs incurred in Kenya [36]It might not be accurate, considering the variation in healthcare standards in study environments. However, this does not affect the trend analysis of incremental costs incurred in different operational configurations.

The operational model in this report assumes that 8% of samples collected for testing each day in the population (derived from the Epidemiological model) enter the WGS network and are exogenously determined. Also, uncertainty is assumed to account for which collection site the samples are collected. We also calibrate the model to estimate the sequencing capacity, which is a close but inaccurate representation of reality. With the help of granular data, one can formulate the exact proportion of samples collected for sequencing in combination with independent modelling of each decentralized GSL with respective sequencing capacities. We have used exogenous variant emergence times and variant characteristics. Researchers can use the probabilities associated with variant mutation and estimate the effectiveness of the WGS network with other public interventions.

## Data Availability

All data and simulations produced in the present study are available upon reasonable request to the authors.

## Funding

This work was supported by the Bill and Melinda Gates Foundation (BMGF) (Grant ID: INV-003239)

## Acknowledgements

The authors are grateful to clinical, laboratory, public health, and policy experts from institutions including the Centre for DNA Fingerprinting and Diagnostics, the Centre for Cellular & Molecular Biology-Hyderabad, Government Medical College Patiala, Rajiv Gandhi Medical College-Thane, the Public Health Foundation of India, Ashoka University, the Integrated Disease Surveillance Progam Office, and PATH Foundation for sharing their time and insights during the development of this study.

Their input helped the authors better understand the laboratory workflow for genomic sequencing, operational processes at sample collection and sentinel surveillance sites, the establishment and functioning of sequencing laboratories, and the use of genomic surveillance evidence to inform public health decision-making.

## Author contributions

SD and SKD conceptualized the study. MJ and KNAR conducted the analysis, and NA, SD and SKD validated the findings. MJ wrote the first draft of the manuscript, and all authors contributed to the final draft.

## Competing Financial Interests

A.M. and JG are the Center for Global Development employees, which funded this research. Authors MJ, KNAR, SD, and SKD received a research grant from the Center for Global Development during this research. The authors declare that no other competing interests exist.

