## Supplementary Material for "Framework to estimate the cost-effectiveness of the Genome Sequencing-based surveillance network: an integrated operational model-epidemiological model approach"

### 1 Epidemiological Model

#### 1.1 Model Description

As shown in **Figure 1**, initially, the population ($S_{0}$) is susceptible to either variant, while the population with prior infection from variant *y* ($S_{x,y}$ )are susceptible to variant *x*. **Figure 1** shows four different transmission pathways ($T_{x,y}$ ) of pathogen in population, where ‘*x*’ represents the current variant, ‘*y*’ represents the variant of previous infection and *y* =0 represents no prior infection.

In the transmission path, $T_{1,0}$, susceptible population $S_{0}$ moves to $E_{1,0}$ at the rate determined by the proportion of infectious individuals of variant 1 in the interacting population ($i_{p,t}^{1}$) and the effective transmission rate (β). Exposed individuals ($E_{1,0}$) turn infectious after the latency period of (δ^-1^) days.

Exposed individuals become infectious, with the proportion of asymptomatic ($I_{1,0,t}^{A}$), severely ill $(I_{1,0,t}^{S})$,and moderately symptomatic $(I_{1,0,t}^{M})$ being $\alpha$, $\alpha_{s}$ and $\left( 1-\alpha-\alpha_{s} \right)$ respectively. Asymptomatic ($I_{1,0,t}^{A}$) and moderately symptomatic ($I_{1,0,t}^{M}$) individuals recover after $(\gamma_{a})$^-1^ days and $(\gamma_{m})$^-1^ days respectively. Severely ill individuals who do not have access to hospital ($\left( 1-\alpha_{h} \right)(I_{1,0,t}^{S}$)) recover after $(\gamma_{\mathrm{nh}})$^-1^ days or die after $(\mu_{\mathrm{nh}})$ ^-1^ days. Severely ill individuals with access to hospital ($\left( \alpha_{h} \right)(I_{1,0,t}^{S}$)) move to hospital after $\lambda_{h}^{-1}$days and recover after $(\gamma_{h})$^-1^ days or die after $(\mu_{h})$ ^-1^ days. Recovered individuals ($R_{1,0}$) lose immunity in $(i_{l}^{1})$^-1^ days and become partially susceptible ($S_{2,1}$) to variant 2.

Infection spreads similarly in all transmission paths with the transmission parameter of variant 2 transmission ( $T_{2,0}$ and $T_{2,1}$ ) being the product of variant 1 transmission and its respective multiplication factor of $m_{z}$. As in [1], we model the loss of immunity and recovered individuals having reduced susceptibility of $r_{x,y}^{s}$ compared to a baseline of unity.

We augment the SEIHRD model by further dividing each disease state compartment based on individuals' test result status. In each disease compartment $C$, $C^{L}$ denotes the number of individuals eligible for testing, i.e., individuals who have not been tested or have obtained the results of all prior tests while $C^{TW}$denotes the individuals who have been given samples for testing and are waiting for their test results To model an individual’s disease state changing when waiting for test results, we use separate compartments for individuals tested after becoming infectious and waiting for test results (**Supplementary Figure 1** illustrates this for the Severely -ill infectious compartments of individuals infected with variant 1 and no prior infection). This is necessary as a sample collected from an individual in the exposed state, who becomes infectious while waiting for the test result, will return a negative result even after they become infectious. The number of tests and test results is a key input for the operational model to determine the number of samples eligible to be sequenced and estimate the cost of sequencing and time to implement any public health interventions. All the compartments of epidemiological model with their description are listed in **Supplementary Table 1**.

Except $S_{0}$, all the compartments are represented as $X_{a,b}^{Y}$ , where X is one of the disease states S, E, I^A^, I^M^, I^S^, or R, ‘a’ represents the variant of the current infection(1 for variant 1 infection and 2 for variant 2 infection), ‘b’ represents the variant of previous infection(‘1’ for variant 1 infection, ‘2’ for variant 2 infection and 0 for no previous infection) and Y represents status of test results.

Supplementary Table 1: Epidemiological model compartment description

| **Compartment**  **Symbol** | **Description** |
| --- | --- |
| $i_{x,t}^{p}$ | The proportion of variant X infectious individuals in the total interacting  population at time t, where x ϵ (1,2) |
| $S_{0,t}^{L}$ | The population susceptible to variant 1 and variant 2, which is eligible for testing, i.e., they are either untested or have received results of all previous tests till time t |
| $S_{0,t}^{TW}$ | The population susceptible to variant 1 and variant 2 at time t, which is waiting  for their test results |
| $S_{a,b,t}^{L}$ | The population susceptible to variant ‘a’ and previously infected by variant ‘b’ at time t, which is eligible for testing, i.e., they are either untested or have received results of all previous tests till time t |
| $S_{a,b,t}^{TW}$ | The population susceptible to variant ‘a’ and previously infected by variant ‘b’ at time t, who are waiting for their test results |
| $E_{a,b,t}^{L}$ | The population exposed to variant ‘a’ at time t and previously infected by variant ‘b’, who are eligible for testing, i.e., they are either untested or have received results of all previous tests till time t |
| $E_{a,b,t}^{TW}$ | The population exposed to variant ‘a’ at time t and previously infected by variant ‘b’, who are waiting for their test results |
| $I_{a,b,t}^{A,L}$ | The population of infectious asymptomatic individuals at time t, infected by variant ‘a’ at time t and previously infected by variant ‘b’, who are eligible for testing, i.e., they are either untested or have received results of all previous tests till time t |
| $I_{a,b,t}^{A,ETW}$ | The population of infectious asymptomatic individuals at time t, infected by variant ‘a’ at time t and previously infected by variant ‘b’, who are waiting for the result of the test taken in the exposed state |
| $I_{a,b,t}^{A,TW}$ | The population of infectious asymptomatic at time t, infected by variant ‘a’ at time t and previously infected by variant ‘b’, who are waiting for the results of tests taken in the infectious state |
| $I_{a,b,t}^{A,DN}$ | The population of infectious asymptomatic at time t, infected by variant ‘a’ at time t and previously infected by variant ‘b’, who are diagnosed, and not quarantined |
| $I_{a,b,t}^{M,L}$ | The population of moderately symptomatic infectious individuals at time t, infected by variant ‘a’ at time t and previously infected by variant ‘b’, who are eligible for testing, i.e., they are either untested or have received results of all previous tests till time t |
| $I_{a,b,t}^{M,ETW}$ | The population of moderately symptomatic infectious individuals at time t, infected by variant ‘a’ at time t and previously infected by variant ‘b’, who are waiting for the result of the test taken in the exposed state |
| $I_{a,b,t}^{M,TW}$ | The population of moderately symptomatic infectious at time t, infected by variant ‘a’ at time t and previously infected by variant ‘b’, who are waiting for the results of tests taken in the infectious state |
| $I_{a,b,t}^{M,DN}$ | The population of moderately symptomatic infectious at time t, infected by variant ‘a’ at time t and previously infected by variant ‘b’, who are diagnosed, and not quarantined |
| $I_{a,b,t}^{S,L}$ | The population of infectious symptomatic individuals at time t, infected by variant ‘a’ at time t and previously infected by variant ‘b’, who are eligible for testing, i.e., they are either untested or have received results of all previous tests till time t |
| $I_{a,b,t}^{S,ETW}$ | The population of infectious symptomatic individuals at time t, infected by variant ‘a’ at time t and previously infected by variant ‘b’, who are waiting for the result of the test taken in the exposed state |
| $I_{a,b,t}^{S,TW}$ | The population of infectious symptomatic at time t, infected by variant ‘a’ at time t and previously infected by variant ‘b’, who are waiting for the results of tests taken in the infectious state |
| $I_{a,b,t}^{S,DN}$ | The population of infectious symptomatic at time t, infected by variant ‘a’ at time t and previously infected by variant ‘b’, diagnosed, and not quarantined |
| $I_{a,b,t}^{A,DQ}$ | The population of infectious asymptomatic at time t, infected by variant ‘a’ at time t and previously infected by variant ‘b’ and quarantined after diagnosis. |
| $I_{a,b,t}^{M,DQ}$ | The population of moderately symptomatic infectious at time t, infected by variant ‘a’ at time t and previously infected by variant ‘b’ and quarantined after diagnosis. |
| $I_{a,b,t}^{S,DQ}$ | The population of infectious symptomatic at time t, infected by variant ‘a’ at time t and previously infected by variant ‘b’ and quarantined after diagnosis. |
| $H_{a,0,t}$ | The population of hospitalized individuals at time t, infected by variant ‘a’ at time t and previously uninfected. |
| $H_{a,b,t}$ | The population of hospitalized individuals at time t, infected by variant ‘a’ at time t and previously infected by variant ‘b’. |
| $R_{a,b,t}^{IA,ETW}$ | The population recovered from being infectious asymptomatic at time t, infected by variant ‘a’ at time t and previously infected by variant ‘b’ and is waiting for the result of the test taken in the exposed state |
| $R_{a,b,t}^{IA,TW}$ | The population recovered from being infectious asymptomatic at time t,  infected by variant ‘a’ at time t and previously infected by variant ‘b’ and is waiting for the result of the test taken in the infectious state |
| $R_{a,b,t}^{IS,ETW}$ | The population recovered from being infectious symptomatic at time t, infected.  by variant x and is waiting for the result of the test taken in the infectious state |
| $R_{a,b,t}^{IS,TW}$ | The population recovered from being infectious symptomatic at time t, infected  by variant x and is waiting for the result of the test taken in the infectious state |
| $R_{a,b,t}^{N}$ | The recovered population at time t, infected by variant ‘a’ at time t and previously infected by variant ‘b’ and has obtained negative results of the test given in the exposed state after recovering |
| $R_{a,b,t}^{U}$ | The recovered population at time t, infected by variant ‘a’ at time t and previously infected by variant ‘b’ and diagnosed positive while in the infectious state. |
| $R_{a,b,t}^{P}$ | The recovered population at time t, infected by variant ‘a’ at time t and previously infected by variant ‘b’ and obtained positive results after recovering |
| $R_{a,b,t}^{TW}$ | The recovered population at time t, infected by variant x who are waiting for the results of tests taken in the recovered state |
| $R_{a,b,t}^{D}$ | The recovered population at time t, infected by variant ‘a’ at time t and previously infected by variant ‘b’ and has obtained results of the test taken in the recovered state |
| $D_{a,b,t}^{IS,ETW}$ | The deceased population at time t, infected by variant ‘a’ at time t and previously infected by variant ‘b’ and the results of tests taken in the exposed state are awaited |
| $D_{a,b,t}^{IS,TW}$ | The deceased population at time t, infected by variant ‘a’ at time t and previously infected by variant ‘b’ and the results of tests taken in the infectious state are awaited |
| $D_{a,b,t}$ | The deceased population at time t, infected by variant ‘a’ at time t and previously infected by variant ‘b’ and no test results are awaited |

#### 1.2 Epidemiological Model equations

Equations governing the population of every epidemiological model compartment at time *t* are listed below:

**Proportion of infectious individuals**

**Sum of asymptomatic infectious individuals**

$$\sum I_{x,t}^{A}=\left( I_{x,b,t}^{A,L}+I_{x,b,t}^{A,ETW}+I_{x,b,t}^{A,TW}+I_{x,b,t}^{A,DN} \right)where (x,b)\epsilon(\left( 1,0 \right),\left( 1,2 \right),\left( 2,0 \right),\left( 2,1 \right))$$

**Sum of moderately symptomatic infectious individuals**

$$\sum I_{x,t}^{M}=\left( I_{x,b,t}^{M,L}+I_{x,b,t}^{M,ETW}+I_{x,b,t}^{M,TW}+I_{x,b,t}^{M,DN} \right)where (x,b)\epsilon(\left( 1,0 \right),\left( 1,2 \right),\left( 2,0 \right),\left( 2,1 \right))$$

**Sum of Symptomatic infectious individuals:**

$$\sum I_{x,t}^{S}=\left( I_{x,b,t}^{S,L}+I_{x,b,t}^{S,ETW}+I_{x,b,t}^{S,TW}+I_{x,b,t}^{S,DN} \right)where (x,b)\epsilon(\left( 1,0 \right),\left( 1,2 \right),\left( 2,0 \right),(2,1))$$

**Sum of total number of dead individuals:**

$$\sum D_{t}={(D}_{x,y,t}^{IS,ETW}+D_{x,y,t}^{IS,TW}+D_{x,y,t}) where (x,y)\epsilon(\left( 1,0 \right),\left( 1,2 \right),\left( 2,0 \right),\left( 2,1 \right))$$

**Sum of total number of quarantine individuals:**

$$\sum Q_{t}=(I_{x,y,t}^{S,DQ}+I_{x,y,t}^{A,DQ}+I_{x,y,t}^{M,DQ}) where (x,y)\epsilon(\left( 1,0 \right),\left( 1,2 \right),\left( 2,0 \right),\left( 2,1 \right))$$

**Total number of hospitalized individuals**

$$\sum H_{t}\boldsymbol{=(}H_{1,0,t}\boldsymbol{+}H_{1,2,t}\boldsymbol{+}H_{2,0,t}\boldsymbol{+}H_{2,1,t}\boldsymbol{)}$$

**Proportion of variant 1 infectious individuals**

$$i_{1,t}^{p}=\frac{((\kappa*{\Sigma I}_{1,t}^{A}) + (\Sigma I_{1,t}^{M}) + (\Sigma I_{1,t}^{S}))}{n-(\Sigma D_{t}+\Sigma Q_{t}+\Sigma H_{t})}$$

**Proportion of variant 2 infectious individuals**

$$i_{2,t}^{p}=\frac{\left( \left( \kappa*{\Sigma I}_{2,t}^{A} \right) + \left( \Sigma I_{2,t}^{M} \right)+ \left( \Sigma I_{2,t}^{S} \right) \right)}{n-(\Sigma D_{t}+\Sigma Q_{t}+\Sigma H_{t})}$$

**Mortality and recovery rate of hospitalized and non-hospitalized severe individuals**

$$\gamma_{h}=h_{rate}*(1-p_{m})$$

$$\mu_{h}\boldsymbol{=}h_{rate}*(p_{m})$$

$$\gamma_{nh}^{-1}=\lambda_{h}^{-1}+(h_{factor}*\gamma_{h}^{-1})$$

$$\mu_{nh}^{-1}=\lambda_{h}^{-1}+(\mu_{h}^{-1}/h_{factor})$$

**Susceptible Compartment Transmission Dynamics**

$S_{0,t+1}^{L}=S_{0,t}^{L}- (\varphi_{s}*S_{0,t}^{L})+ (\omega*S_{0,t}^{TW})- (i_{2,t}^{p}*m_{\beta}*\beta*S_{0,t}^{L})- (i_{1,t}^{p}$*β*$S_{0,t}^{L})$

$S_{0,t+1}^{TW}=S_{0,t}^{TW}- \left( i_{1,t}^{p}*\beta*S_{0,t}^{TW} \right)-\left( \omega*S_{0,t}^{TW} \right)+\left( \varphi_{s}*S_{0,t}^{L} \right)-\left( i_{2,t}^{p}*m_{\beta}*\beta*S_{0,t}^{TW} \right)$

$S_{x,y,t+1}^{L}=S_{x,y,t}^{L}- \left( \varphi_{s}*S_{x,y,t}^{L} \right)+ \left( \omega*S_{x,y,t}^{TW} \right)- \left( {r_{x,y}^{s}*i}_{x,t}^{p}*c*S_{x,y,t}^{L} \right)+\left( i_{y}^{L}*{(R}_{y,0,t}^{N}+R_{y,0,t}^{U}+R_{y,0,t}^{P}+R_{y,0,t}^{D}) \right), where \left( x,y \right)\epsilon\left( \left( 1,2 \right),\left( 2,1 \right) \right) and c=\left\{ \begin{aligned} \beta, &x,y=1,2 \\ m_{\beta}*\beta, &x,y=2,1 \end{aligned} \right.$

$$S_{x,y,t+1}^{TW}=S_{x,y,t}^{TW}- \left( r_{x,y}^{s}*i_{1,t}^{p}*c*S_{x,y,t}^{TW} \right)-\left( \omega*S_{x,y,t}^{TW} \right)+\left( \varphi_{s}*S_{x,y,t}^{L} \right)+ +\left( i_{y}^{L}*\left( R_{y,0,t}^{IS,ETW}+R_{y,0,t}^{IA,ETW}+R_{y,0,t}^{IS,TW}+R_{y,0,t}^{IA,TW}+R_{y,0,t}^{TW}+ \right) \right), where \left( x,y \right)\epsilon\left( \left( 1,2 \right),\left( 2,1 \right) \right)and c=\left\{ \begin{aligned} \beta, &x,y=1,2 \\ m_{\beta}*\beta, &x,y=2,1 \end{aligned} \right.$$

**Exposed Compartment Transmission Dynamics**

$E_{x,0,t+1}^{L} =E_{x,0,t}^{L}+\left( i_{x,t}^{p}*c*S_{0,t}^{L} \right)+\left( \omega* E_{x,0,t}^{TW} \right)-\left( z*E_{x,0,t}^{L} \right)-\left( \varphi_{e}*E_{x,0,t}^{L} \right), where x \epsilon(1,2), b=\left\{ \begin{aligned} \beta, &x=1 \\ m_{\beta}*\beta, &x=2 \end{aligned} ,and z=\left\{ \begin{aligned} \delta, &x=1 \\ m_{\delta}*\delta, &x=2 \end{aligned} \right. \right.$

$E_{x,0,t+1}^{TW}=E_{x,0,t}^{TW}+\left( \varphi_{e}*E_{x,0,t}^{L} \right)+\left( i_{x,t}^{p}*c*S_{0,t}^{TW} \right)-\left( \omega*E_{x,0,t}^{TW} \right)-\left( z*E_{x,0,t}^{TW} \right), where x\epsilon\left( 1,2 \right), c=\left\{ \begin{aligned} \beta, &x=1 \\ m_{\beta}*\beta, &x=2 \end{aligned} , and z=\left\{ \begin{aligned} \delta, &x=1 \\ m_{\delta}*\delta, &x=2 \end{aligned} \right. \right.$

$E_{x,y,t+1}^{L} =E_{x,y,t}^{L}+\left( {r_{x,y}^{s}*i}_{x,t}^{p}*c*S_{x,y,t}^{L} \right)+\left( \omega* E_{x,y,t}^{TW} \right)-\left( z*E_{x,y,t}^{L} \right)-\left( \varphi_{e}*E_{x,y,t}^{L} \right), where \left( x,y \right)\epsilon\left( \left( 1,2 \right),\left( 2,1 \right) \right), c=\left\{ \begin{aligned} \beta, &x,y=1,2 \\ \beta*m\beta, &x,y=2,1 \end{aligned} ,and z=\left\{ \begin{aligned} \delta, &x,y=1,2 \\ m_{\delta}*\delta, &x,y=2,1 \end{aligned} \right. \right.$

$E_{x,y,t+1}^{TW}=E_{x,y,t}^{TW}+\left( \varphi_{e}*E_{x,y,t}^{L} \right)+\left( r_{x,y}^{s}*i_{1,t}^{p}*c*S_{x,y,t}^{TW} \right)-\left( \omega*E_{x,y,t}^{TW} \right)-\left( z*E_{x,y,t}^{TW} \right), where (x,y)\epsilon((1,2),(2,1)), c=\left\{ \begin{aligned} \beta, &x,y=1,2 \\ m_{\beta}*\beta, &x,y=2,1 \end{aligned} , and z=\left\{ \begin{aligned} \delta, &x,y=1,2 \\ m_{\delta}*\delta, &x,y=2,1 \end{aligned} \right. \right.$

**Infectious-asymptomatic transmission equations**

$$I_{x,y,t+1}^{A,L}= I_{x,y,t}^{A,L}+\left( z*l^{a}*E_{x,y,t}^{L} \right)+\left( \omega*I_{x,y,t}^{A,ETW} \right)- \left( \varphi_{i,a}*I_{x,y,t}^{A,L} \right)- \left( g^{a}*I_{x,y,t}^{A,L} \right), where \left( x,y \right)\epsilon\left( \left( 1,0 \right),\left( 2,0 \right),\left( 1,2 \right),\left( 2,1 \right) \right), g^{a}=\left\{ \begin{aligned} \gamma_{a}, &x=1 \\ m_{\gamma,a}*\gamma_{a}, &x=2 \end{aligned} \right., l^{a}=\left\{ \begin{aligned} \alpha, &x=1 \\ m_{\alpha}*\alpha, &x=2 \end{aligned} \right.and z=\left\{ \begin{aligned} \delta, &x=1 \\ m_{\delta}*\delta, &x=2 \end{aligned} \right.$$

$I_{x,y,t+1}^{A,ETW}= I_{x,y,t}^{A,ETW}+\left( z*l^{a}*E_{x,y,t}^{TW} \right)-\left( \omega*I_{x,y,t}^{A,ETW} \right)- \left( g^{a}*I_{x,y,t}^{A,ETW} \right),where \left( x,y \right)\epsilon\left( \left( 1,0 \right),\left( 2,0 \right),\left( 1,2 \right),\left( 2,1 \right) \right), l^{a}=\left\{ \begin{aligned} \alpha, &x=1 \\ m_{\alpha}*\alpha, &x=2 \end{aligned} \right.and z=\left\{ \begin{aligned} \delta, &x,y=1,2 \\ m_{\delta}*\delta, &x,y=2,1 \end{aligned} \right. , g^{a}=\left\{ \begin{aligned} \gamma_{a}, &x=1 \\ m_{\gamma,a}*\gamma_{a}, &x=2 \end{aligned} \right.$

$$I_{x,y,t+1}^{A,TW}= I_{x,y,t}^{A,TW}+\left( \varphi_{i,a}*I_{x,y,t}^{A,L} \right)- \left( \omega*I_{x,y,t}^{A,TW} \right)- \left( g^{a}*I_{x,y,t}^{A,TW} \right), where \left( x,y \right)\epsilon\left( \left( 1,0 \right),\left( 2,0 \right),\left( 1,2 \right),\left( 2,1 \right) \right)and g^{a}=\left\{ \begin{aligned} \gamma_{a}, &x=1 \\ m_{\gamma,a}*\gamma_{a}, &x=2 \end{aligned} \right.$$

$I_{x,y,t+1}^{A,DN}= I_{x,y,t}^{A,DN} + ((1 - q)*\omega*I_{x,y,t}^{A,TW}) -(g^{a}*I_{x,y,t}^{A,DN}), where (x,y)\epsilon((1,0),(2,0),(1,2),(2,1)) and g^{a}=\left\{ \begin{aligned} \gamma_{a}, &x=1 \\ m_{\gamma,a}*\gamma_{a}, &x=2 \end{aligned} \right.$

**Infectious-moderately symptomatic transmission equations**

$$I_{x,y,t+1}^{M,L}= I_{x,y,t}^{M,L}+\left( z*l^{m}*E_{x,y,t}^{L} \right)+\left( \omega*I_{x,y,t}^{M,ETW} \right)- \left( \varphi_{i,m}*I_{x,y,t}^{M,L} \right)- \left( g^{m}*I_{x,y,t}^{M,L} \right), where \left( x,y \right)\epsilon\left( \left( 1,0 \right),\left( 2,0 \right),\left( 1,2 \right),\left( 2,1 \right) \right),g^{m}=\left\{ \begin{aligned} \gamma_{m}, &x=1 \\ m_{\gamma,m}*\gamma_{m}, &x=2 \end{aligned} \right., l^{m}=\left\{ \begin{aligned} {1-(\alpha+\alpha}_{s}), &x=1 \\ {1-(m_{\alpha}\alpha+m_{\alpha,s}\alpha}_{s}), &x=2 \end{aligned} \right.and z=\left\{ \begin{aligned} \delta, &x=1 \\ m_{\delta}*\delta, &x=2 \end{aligned} \right.$$

$I_{x,y,t+1}^{M,ETW}=I_{x,y,t}^{M,ETW}+\left( z* l^{m}*E_{x,y,t}^{TW} \right)-\left( \omega*I_{x,y,t}^{M,ETW} \right)-\left( g^{m}*I_{x,y,t}^{M,ETW} \right), where \left( x,y \right)\epsilon\left( \left( 1,0 \right),\left( 2,0 \right),\left( 1,2 \right),\left( 2,1 \right) \right), l^{m}=\left\{ \begin{aligned} {1-(\alpha+\alpha}_{s}), &x=1 \\ {1-(m_{\alpha}\alpha+m_{\alpha,s}\alpha}_{s}), &x=2 \end{aligned} \right., z=\left\{ \begin{aligned} \delta, &x,y=1,2 \\ m_{\delta}*\delta, &x,y=2,1 \end{aligned} \right. and g^{m}=\left\{ \begin{aligned} \gamma_{m}, &x=1 \\ m_{\gamma,m}*\gamma_{m}, &x=2 \end{aligned} \right.$

$$I_{x,y,t+1}^{M,TW}= I_{x,y,t}^{M,TW}+\left( \varphi_{i,m}*I_{x,y,t}^{M,L} \right)- \left( *I_{x,y,t}^{M,TW} \right)- \left( g^{m}*I_{x,y,t}^{M,TW} \right), where (x,y)\epsilon((1,0),(2,0),(1,2),(2,1)) and g^{m}=\left\{ \begin{aligned} \gamma_{m}, &x=1 \\ m_{\gamma,m}*\gamma_{m}, &x=2 \end{aligned} \right.$$

$$I_{x,y,t+1}^{M,DN}=I_{x,y,t}^{M,DN}+\left( \left( 1-q \right)*\omega*I_{x,y,t}^{M,TW} \right)-\left( g^{m}*I_{x,y,t}^{M,DN} \right), where \left( x,y \right)\epsilon\left( \left( 1,0 \right),\left( 2,0 \right),\left( 1,2 \right),\left( 2,1 \right) \right) and g^{m}=\left\{ \begin{aligned} \gamma_{m}, &x=1 \\ m_{\gamma,m}*\gamma_{m}, &x=2 \end{aligned} \right.$$

**Infectious-severe symptomatic transmission equations**

$$I_{x,y,t+1}^{S,L}=I_{x,y,t}^{S,L}+\left( z*l^{s}*E_{x,y,t}^{L} \right)+\left( \omega*I_{x,y,t}^{S,ETW} \right)-\left( \varphi_{i,s}*I_{x,y,t}^{S,L} \right)-\left( g^{nh}*I_{1,b,t}^{S,L} \right)-\left( h^{r}*I_{1,b,t}^{S,L} \right) - \left( d^{nh}*I_{1,b,t}^{S,L} \right), where \left( x,y \right)\epsilon\left( \left( 1,0 \right),\left( 2,0 \right),\left( 1,2 \right),\left( 2,1 \right) \right),g^{nh}=\left\{ \begin{aligned} \gamma_{nh}, &x=1 \\ m_{\gamma,nh}*\gamma_{nh}, &x=2 \end{aligned} \right., l^{s}=\left\{ \begin{aligned} \alpha_{s}, &x=1 \\ m_{\alpha,s}*\alpha_{s}, &x=2 \end{aligned} \right., z=\left\{ \begin{aligned} \delta, &x=1 \\ m_{\delta}*\delta, &x=2 \end{aligned} \right.,h^{r}=\left\{ \begin{aligned} \lambda, &x=1 \\ m_{\lambda}*\lambda, &x=2 \end{aligned} \right.and d^{nh}=\left\{ \begin{aligned} \mu_{nh}, &x=1 \\ m_{\mu,nh}*\mu_{nh}, &x=2 \end{aligned} \right.$$

$I_{x,y,t+1}^{S,ETW}= I_{x,y,t}^{S,ETW}+\left( z*l^{s}*E_{x,y,t}^{TW} \right)-\left( \omega*I_{x,y,t}^{S,ETW} \right)- \left( g^{nh}*I_{x,y,t}^{S,ETW} \right)-\left( h^{r}*I_{1,b,t}^{S,ETW} \right)-\left( d^{nh}*I_{x,y,t}^{S,ETW} \right), where \left( x,y \right)\epsilon\left( \left( 1,0 \right),\left( 2,0 \right),\left( 1,2 \right),\left( 2,1 \right) \right), g^{nh}=\left\{ \begin{aligned} \gamma_{nh}, &x=1 \\ m_{\gamma,nh}*\gamma_{nh}, &x=2 \end{aligned} \right., l^{s}=\left\{ \begin{aligned} \alpha_{s}, &x=1 \\ m_{\alpha,s}*\alpha_{s}, &x=2 \end{aligned} \right., z=\left\{ \begin{aligned} \delta, &x=1 \\ m_{\delta}*\delta, &x=2 \end{aligned} \right., h^{r}=\left\{ \begin{aligned} \lambda, &x=1 \\ m_{\lambda}*\lambda, &x=2 \end{aligned} \right.and d^{nh}=\left\{ \begin{aligned} \mu_{nh}, &x=1 \\ m_{\mu,nh}*\mu_{nh}, &x=2 \end{aligned} \right.$

$$I_{x,y,t+1}^{S,TW}=I_{x,y,t}^{S,TW}+\left( \varphi_{i,s}*I_{x,y,t}^{S,L} \right)-\left( \omega*I_{x,y,t}^{S,TW} \right)-\left( g^{nh}*I_{x,y,t}^{S,TW} \right)-\left( h^{r}*I_{1,b,t}^{S,TW} \right)-\left( d^{nh}*I_{x,y,t}^{S,TW} \right), where \left( x,y \right)\epsilon\left( \left( 1,0 \right),\left( 2,0 \right),\left( 1,2 \right),\left( 2,1 \right) \right),g^{nh}=\left\{ \begin{aligned} \gamma_{nh}, &x=1 \\ m_{\gamma,nh}*\gamma_{nh}, &x=2 \end{aligned} \right., h^{r}=\left\{ \begin{aligned} \lambda, &x=1 \\ m_{\lambda}*\lambda, &x=2 \end{aligned} \right.and d^{nh}=\left\{ \begin{aligned} \mu_{nh}, &x=1 \\ m_{\mu,nh}*\mu_{nh}, &x=2 \end{aligned} \right.$$

$$I_{x,y,t+1}^{S,DN}=I_{x,y,t}^{S,DN}+\left( \left( 1-q \right)*\omega*I_{x,y,t}^{S,TW} \right) -\left( g^{nh}*I_{x,y,t}^{S,DN} \right)-\left( h^{r}*I_{1,b,t}^{S,DN} \right) - \left( d^{nh}*I_{x,y,t}^{S,DN} \right), where \left( x,y \right)\epsilon\left( \left( 1,0 \right),\left( 2,0 \right),\left( 1,2 \right),\left( 2,1 \right) \right), g^{nh} =\left\{ \begin{aligned} \gamma_{nh}, &x=1 \\ m_{\gamma,nh}*\gamma_{nh}, &x=2 \end{aligned} \right.,h^{r}=\left\{ \begin{aligned} \lambda, &x=1 \\ m_{\lambda}*\lambda, &x=2 \end{aligned} \right.and d^{nh}= \begin{aligned} \mu_{nh}, &x=1 \\ m_{\mu,nh}*\mu_{nh}, &x=2 \end{aligned}$$

**Quarantine compartment equations**

$$I_{x,y,t+1}^{A,DQ}= I_{x,y,t}^{A,DQ} + ( q*\omega*I_{x,y,t}^{A,TW}) -(g^{a}*I_{x,y,t}^{A,DQ}), where (x,y)\epsilon((1,0),(2,0),(1,2),(2,1)) and g^{a}=\left\{ \begin{aligned} \gamma_{a}, &x=1 \\ m_{\gamma,a}*\gamma_{a}, &x=2 \end{aligned} \right.$$

$$I_{x,y,t+1}^{S,DQ}=I_{x,y,t}^{S,DQ}+\left( q*\omega*I_{x,y,t}^{S,TW} \right)-\left( g^{nh}*I_{x,y,t}^{S,DQ} \right)-\left( h^{r}*I_{x,y,t}^{S,DQ} \right)-\left( d^{nh}*I_{x,y,t}^{S,DQ} \right), where \left( x,y \right)\epsilon\left( \left( 1,0 \right),\left( 2,0 \right),\left( 1,2 \right),\left( 2,1 \right) \right), g^{nh}=\left\{ \begin{aligned} \gamma_{nh}, &x=1 \\ m_{\gamma,nh}*\gamma_{nh}, &x=2 \end{aligned} \right.,h^{r}=\left\{ \begin{aligned} \lambda, &x=1 \\ m_{\lambda}*\lambda, &x=2 \end{aligned} \right.,and d^{nh}=\left\{ \begin{aligned} \mu_{nh}, &x=1 \\ m_{\mu,nh}*\mu_{nh}, &x=2 \end{aligned} \right.$$

$I_{x,y,t+1}^{M,DQ} = I_{x,y,t}^{M,DQ}+\left( q*\omega*I_{x,y,t}^{M,TW} \right)-\left( g^{m}*I_{x,y,t}^{M,DQ} \right), where (x,y)\epsilon((1,0),(2,0),(1,2),(2,1)) and g^{m}=\left\{ \begin{aligned} \gamma_{m}, &x=1 \\ m_{\gamma,m}*\gamma_{m}, &x=2 \end{aligned} \right.$

**Hospitalization Compartment Equation**

$$H_{x,y,t+1}= H_{x,y,t}+h^{r}*\left( I_{x,y,t}^{S,L}+I_{x,y,t}^{S,ETW}+I_{x,y,t}^{S,TW}+I_{x,y,t}^{S,DN}+I_{x,y,t}^{S,DQ} \right)-\left( g^{h}*H_{x,y,t} \right)-\left( d^{h}*H_{x,y,t} \right), where \left( x,y \right)\epsilon\left( \left( 1,0 \right),\left( 2,0 \right),\left( 1,2 \right),\left( 2,1 \right) \right), g^{h} =\left\{ \begin{aligned} \gamma_{h}, &x=1 \\ m_{\gamma,h}*\gamma_{h}, &x=2 \end{aligned} \right.,h^{r}=\left\{ \begin{aligned} \lambda, &x=1 \\ m_{\lambda}*\lambda, &x=2 \end{aligned} \right., and d^{h}=\left\{ \begin{aligned} \mu_{h}, &x=1 \\ m_{\mu,h}*\mu_{h}, &x=2 \end{aligned} \right.$$

**Recovered Compartment equations**

$$R_{x,0,t+1}^{IA,ETW}= R_{x,0,t}^{IA,ETW}+\left( g^{a}*I_{x,0,t}^{A,ETW} \right)-\left( \omega*R_{x,0,t}^{IA,ETW} \right)-\left( i_{x}^{L}*R_{x,0,t}^{IA,ETW} \right),where x\epsilon(1,2) andg^{a}=\left\{ \begin{aligned} \gamma_{a}, &x=1 \\ m_{\gamma,a}*\gamma_{a}, &x=2 \end{aligned} \right.$$

$$R_{x,0,t+1}^{IS,ETW}= R_{x,0,t}^{IS,ETW}+\left( g^{nh}*I_{x,0,t}^{S,ETW} \right)+\left( g^{m}*I_{x,0,t}^{M,ETW} \right)-\left( \omega*R_{x,0,t}^{IS,ETW} \right)-\left( i_{x}^{L}*R_{x,0,t}^{IS,ETW} \right), where x\epsilon\left( 1,2 \right), g^{m}=\left\{ \begin{aligned} \gamma_{m}, &x=1 \\ m_{\gamma,m}*\gamma_{m}, &x=2 \end{aligned} \right., and g^{nh}=\left\{ \begin{aligned} \gamma_{nh}, &x=1 \\ m_{\gamma,nh}*\gamma_{nh}, &x=2 \end{aligned} \right.$$

$$R_{x,0,t+1}^{IA,TW}= R_{x,0,t}^{IA,TW}+\left( z*I_{x,0,t}^{A,TW} \right)-\left( \omega*R_{x,0,t}^{IA,TW} \right)-\left( i_{x}^{L}*R_{x,0,t}^{IA,TW} \right), where x\epsilon(1,2) and z=\left\{ \begin{aligned} \gamma_{a}, &x=1 \\ m_{\gamma,a}*\gamma_{a}, &x=2 \end{aligned} \right.$$

$$R_{x,0,t+1}^{IS,TW}= R_{x,0,t}^{IS,TW}+\left( g^{nh}*I_{x,0,t}^{S,TW} \right)+\left( g^{m}*I_{x,0,t}^{M,TW} \right)-\left( \omega*R_{x,0,t}^{IS,TW} \right)-\left( i_{x}^{L}*R_{x,0,t}^{IS,TW} \right), where x\epsilon(1,2), g^{m}=\left\{ \begin{aligned} \gamma_{m}, &x=1 \\ m_{\gamma,m}*\gamma_{m}, &x=2 \end{aligned} \right., andg^{nh}=\left\{ \begin{aligned} \gamma_{nh}, &x=1 \\ m_{\gamma,nh}*\gamma_{nh}, &x=2 \end{aligned} \right.$$

$$R_{x,0,t+1}^{N}=R_{x,0,t}^{N}+(\omega*(R_{x,0,t}^{IA,ETW}+R_{x,0,t}^{IS,ETW})) - (\varphi_{r,n} *R_{x,0,t}^{N}) - (i_{x}^{L}*R_{x,0,t}^{N}), where x\epsilon(1,2)$$

$$R_{x,0,t+1}^{L}=R_{x,0,t}^{L} - \left( \varphi_{r,u} *R_{x,0,t}^{L} \right)+\left( g^{a}*\left( I_{x,0,t}^{A,L}+I_{x,0,t}^{A,DN}+I_{x,b,t}^{A,DQ} \right) \right)+ \left( g^{nh}*\left( I_{x,0,t}^{S,L}+I_{x,0,t}^{S,DN}+I_{x,b,t}^{S,DQ} \right) \right)+\left( g^{m}*\left( I_{x,0,t}^{M,L}+I_{x,0,t}^{M,DN}+I_{x,b,t}^{M,DQ} \right) \right)-\left( i_{x}^{L}*R_{x,0,t}^{N} \right), where x\epsilon\left( 1,2 \right), g^{a}=\left\{ \begin{aligned} \gamma_{a}, &x=1 \\ m_{\gamma,a}*\gamma_{a}, &x=2 \end{aligned} \right.,g^{nh}=\left\{ \begin{aligned} \gamma_{nh}, &x=1 \\ m_{\gamma,nh}*\gamma_{nh}, &x=2 \end{aligned} ,and g^{m}=\left\{ \begin{aligned} \gamma_{m}, &x=1 \\ m_{\gamma,m}*\gamma_{m}, &x=2 \end{aligned} \right. \right.$$

$$R_{x,0,t+1}^{P}=R_{x,0,t}^{P}+(\omega*(R_{x,0,t}^{IA,TW}+R_{x,0,t}^{IS,TW})) - (\varphi_{r,p} *R_{x,0,t}^{P}) - (i_{x}^{L}*R_{x,0,t}^{P}), where x\epsilon(1,2)$$

$$R_{x,0,t+1}^{TW}=R_{x,0,t}^{TW}+\left( \varphi r,n *R_{x,0,t}^{N} \right)+\left( \varphi_{r,u} *R_{x,0,t}^{L} \right)+ \left( \varphi_{r,p} *R_{x,0,t}^{P} \right)- \left( \omega*R_{x,0,t}^{TW} \right)- \left( i_{x}^{L}*R_{x,0,t}^{TW} \right), where x\epsilon(1,2)$$

$$R_{x,0,t+1}^{D}=R_{x,0,t}^{D}+\left( \omega*R_{x,0,t}^{TW} \right)- \left( i_{x}^{L}*R_{x,0,t}^{D} \right), where x\epsilon(1,2)$$

$$R_{x,y,t+1}^{IA,ETW}= R_{x,y,t}^{IA,ETW}+\left( g^{a}*I_{x,y,t}^{A,ETW} \right)-\left( \omega*R_{x,y,t}^{IA,ETW} \right), where (x,y)\epsilon((1,2),(2,1)), and g^{a}=\left\{ \begin{aligned} \gamma_{a}, &x=1,2 \\ m_{\gamma,a}*\gamma_{a}, &x=2,1 \end{aligned} \right.$$

$$R_{x,y,t+1}^{IS,ETW}= R_{x,y,t}^{IS,ETW}+\left( g^{nh}*I_{x,y,t}^{S,ETW} \right)+\left( g^{m}*I_{x,y,t}^{M,ETW} \right)-\left( \omega*R_{x,y,t}^{IS,ETW} \right), where \left( x,y \right)\epsilon\left( \left( 1,2 \right),\left( 2,1 \right) \right),g^{m}=\left\{ \begin{aligned} \gamma_{m}, &x=1 \\ m_{\gamma,m}*\gamma_{m}, &x=2 \end{aligned} \right., and g^{nh}=\left\{ \begin{aligned} \gamma_{nh}, &x=1 \\ m_{\gamma,nh}*\gamma_{nh}, &x=2 \end{aligned} \right.$$

$$R_{x,y,t+1}^{IA,TW}= R_{x,y,t}^{IA,TW}+\left( g^{a}*I_{x,y,t}^{A,TW} \right)-\left( \omega*R_{x,y,t}^{IA,TW} \right), where (x,y)\epsilon((1,2),(2,1)), andg^{a}=\left\{ \begin{aligned} \gamma a, &x=1 \\ m_{\gamma,a}*\gamma_{a}, &x=2 \end{aligned} \right.$$

$$R_{x,y,t+1}^{IS,TW}=R_{x,y,t}^{IS,TW}+\left( g^{nh}*I_{x,y,t}^{S,TW} \right)+\left( g^{m}*I_{x,y,t}^{M,TW} \right)-\left( \omega*R_{x,y,t}^{IS,TW} \right), where \left( x,y \right)\epsilon\left( \left( 1,2 \right),\left( 2,1 \right) \right),g^{m}=\left\{ \begin{aligned} \gamma_{m}, &x=1 \\ m_{\gamma,m}*\gamma_{m}, &x=2 \end{aligned} \right. ,and g^{nh}=\left\{ \begin{aligned} \gamma_{nh}, &x=1 \\ m_{\gamma,nh}*\gamma_{nh}, &x=2 \end{aligned} \right.$$

$$R_{x,y,t+1}^{N}=R_{x,y,t}^{N}+\left( \omega*\left( R_{x,y,t}^{IA,ETW}+R_{x,y,t}^{IS,ETW} \right) \right)- \left( \varphi_{r,n} *R_{x,y,t}^{N} \right), where (x,y)\epsilon((1,2),(2,1))$$

$$R_{x,y,t+1}^{L}=R_{x,y,t}^{L} - \left( \varphi_{r,u} *R_{x,y,t}^{L} \right)+\left( g^{a}*\left( I_{x,y,t}^{A,L}+I_{x,y,t}^{A,DN}+I_{x,y,t}^{A,DQ} \right) \right)+ \left( g^{nh}*\left( I_{x,y,t}^{S,L}+I_{x,y,t}^{S,DN}+I_{x,y,t}^{S,DQ} \right) \right)+\left( g^{m}*\left( I_{x,y,t}^{M,L}+I_{x,y,t}^{M,DN}+I_{x,y,t}^{M,DQ} \right) \right), where \left( x,y \right)\epsilon\left( \left( 1,2 \right),\left( 2,1 \right) \right),g^{a}=\left\{ \begin{aligned} \gamma_{a}, &x=1 \\ m_{\gamma,a}*\gamma_{a}, &x=2 \end{aligned} \right.,g^{nh}=\left\{ \begin{aligned} \gamma_{nh}, &x=1 \\ m_{\gamma,nh}*\gamma_{nh}, &x=2 \end{aligned} \right. and g^{m}=\left\{ \begin{aligned} \gamma_{m}, &x=1 \\ m_{\gamma,m}*\gamma_{m}, &x=2 \end{aligned} \right.$$

$$R_{x,y,t+1}^{P}=R_{x,y,t}^{P}+(\omega*(R_{x,y,t}^{IA,TW}+R_{x,y,t}^{IS,TW})) - (\varphi_{r,p} *R_{x,y,t}^{P}), where (x,y)\epsilon((1,2),(2,1))$$

$$R_{x,y,t+1}^{TW}=R_{x,y,t}^{TW}+(\varphi_{r,n} *R_{x,y,t}^{N}) +(\varphi_{r,u} *R_{x,y,t}^{L})+ (\varphi_{r,p} *R_{x,y,t}^{P})- (\omega*R_{x,y,t}^{TW}), where (x,y)\epsilon((1,2),(2,1))$$

$$R_{x,y,t+a}^{D}=R_{x,y,t}^{D}+\left( \omega*R_{x,y,t}^{TW} \right), where (x,y)\epsilon((1,2),(2,1))$$

**Deceased compartment equations**

$$D_{x,y,t+1}= D_{x,y,t}+\left( d^{nh}*\left( I_{x,y,t}^{S,L}+I_{x,y,t}^{S,DN} + I_{x,y,t}^{S,DQ} \right) \right)+\left( d^{h}*H_{x,y,t} \right)+\left( \omega*D_{x,y,t}^{IS,ETW} \right)+\left( \omega*D_{x,y,t}^{IS,TW} \right), where \left( x,y \right)\epsilon\left( \left( 1,0 \right),\left( 2,0 \right),\left( 1,2 \right),\left( 2,1 \right) \right), and d^{nh}=\left\{ \begin{aligned} \mu_{nh}, &x=1 \\ m_{\mu,nh}*\mu_{nh}, &x=2 \end{aligned} \right., and d^{h}=\left\{ \begin{aligned} \mu_{h}, &x=1 \\ m_{\mu,h}*\mu_{h}, &x=2 \end{aligned} \right.$$

$$D_{x,y,t+1}^{IS,ETW} =D_{x,y,t}^{IS,ETW}+\left( d^{nh}*I_{x,y,t}^{S,ETW} \right)-\left( \omega*D_{x,y,t}^{IS,ETW} \right), where \left( x,y \right)\epsilon\left( \left( 1,0 \right),\left( 2,0 \right),\left( 1,2 \right),\left( 2,1 \right) \right),and d^{nh}=\left\{ \begin{aligned} \mu_{nh}, &x=1 \\ m_{\mu,nh}*\mu_{nh}, &x=2 \end{aligned} \right.$$

$$D_{x,y,t+1}^{IS,TW}= D_{x,y,t}^{IS,TW}+\left( d^{nh}*I_{x,y,t}^{S,TW} \right)-\left( \omega*D_{x,y,t}^{IS,TW} \right), where \left( x,y \right)\epsilon\left( \left( 1,0 \right),\left( 2,0 \right),\left( 1,2 \right),\left( 2,1 \right) \right) and d^{nh}=\left\{ \begin{aligned} \mu_{nh}, &x=1 \\ m_{\mu,nh}*\mu_{nh}, &x=2 \end{aligned} \right.$$

#### 1.3 Average Life-year saved

To calculate the average number of life-years saved on averting a death, we used the on the age-wise comparison of COVID deaths [2] (see **Supplementary Table 2**)and the expected life years left of an individual in an age-group from [3] (see **Supplementary Table 3**). The age-groups in [3] is 10 years while the expected life-years are calculated in age- groups of 5 years. To estimate the expected life years left in 10-year age group scale, we use the data from [4] (see **Supplementary Table 4**) which gives the projected population of India in 2021 on 10-year age groups. We assign the weights to the population in the TABLE based on their population percentage in TABLE and calculated the average number of life years saved on saving a death from an age-group as shown in **Supplementary Table 5** column D. The average life-years saved on averting a death is calculated by assigning the weights to an age group in Supplementary Table 2 based on percentage deaths as shown in **Supplementary Table 6**.

Supplementary Table 2: Percentage of all COVID-19 Deaths in an age-group

| **Age group** | **Deaths (%)** |
| --- | --- |
| < 10 | 0.34 |
| 10-20 | 0.31 |
| 20-30 | 1.72 |
| 30-40 | 5.39 |
| 40—50 | 10.82 |
| 50—60 | 21.23 |
| 60—70 | 28.21 |
| 70—80 | 22.17 |
| > 80 | 9.81 |

Supplementary Table 3: Additional life-years left of an individual in an age-group

| **Age Interval (x to x+n)** | **Additional life years left** |
| --- | --- |
| 0-1 | 70 |
| 1-5 | 71.5 |
| 5-10 | 67.8 |
| 10-15 | 63 |
| 15-20 | 58.2 |
| 20-25 | 53.4 |
| 25-30 | 48.7 |
| 30-35 | 44 |
| 35-40 | 39.3 |
| 40-45 | 34.8 |
| 45-50 | 30.3 |
| 50-55 | 26 |
| 55-60 | 22 |
| 60-65 | 18.3 |
| 65-70 | 14.9 |
| 70-75 | 11.9 |
| 75-80 | 9.3 |
| 80-85 | 7 |
| 85+ | 5.3 |

Notes: This table is taken from ‘Abridged Life Tables 2016-2020’ [3] and estimates the average number of additional years person would live if the current mortality trends were to continue.

Supplementary Table 4: Projected population in 2021 different age groups

| **Age Group** | **Total population** |
| --- | --- |
| 0-1 | 23,295 |
| 1-4 | 90978 |
| 5-9 | 1,17,666 |
| 10-14 | 1,18,051 |
| 15-19 | 1,24,282 |
| 20-24 | 1,27,244 |
| 25-29 | 1,19,900 |
| 30-34 | 1,09,575 |
| 35-39 | 98,863 |
| 40-44 | 88,765 |
| 45-49 | 80,107 |
| 50-54 | 69,566 |
| 55-59 | 57,144 |
| 60-64 | 44,544 |
| 65-69 | 34,406 |
| 70-74 | 26,450 |
| 75-79 | 17,563 |
| 80+ | 14,607 |
| All ages | 13,63,006 |

Notes : This table is taken from ‘Report of Technical Analysis Group on population projections’ [4] and represents population in sample size of 13,63,006.

Supplementary Table 5: Average additional life years for age-group in COVID mortality data

| **Age Group**  **A** | **Additional life years left**  **B** | **Proportion of total population**  **C** | **Average additional life years left**  **D** | **Age groups, as per COVID mortality data**  **E** |
| --- | --- | --- | --- | --- |
| 0-1 | 70 | 0.017 | 69.47 | <10 |
| 1-5 | 71.5 | 0.067 |  |  |
| 5-10 | 67.8 | 0.086 |  |  |
| 10-15 | 63 | 0.087 | 60.54 | 10-20 |
| 15-20 | 58.2 | 0.091 |  |  |
| 20-25 | 53.4 | 0.093 | 51.12 | 20-30 |
| 25-30 | 48.7 | 0.088 |  |  |
| 30-35 | 44 | 0.080 | 41.77 | 30-40 |
| 35-40 | 39.3 | 0.073 |  |  |
| 40-45 | 34.8 | 0.065 | 32.67 | 40—50 |
| 45-50 | 30.3 | 0.059 |  |  |
| 50-55 | 26 | 0.051 | 24.20 | 50—60 |
| 55-60 | 22 | 0.042 |  |  |
| 60-65 | 18.3 | 0.033 | 16.82 | 60—70 |
| 65-70 | 14.9 | 0.025 |  |  |
| 70-75 | 11.9 | 0.019 | 10.86 | 70—80 |
| 75-80 | 9.3 | 0.013 |  |  |
| 80-85 | 7 | 0.005 | 6.15 | > 80 |
| 85+ | 5.3 | 0.005 |  |  |

Notes: This table merges the proportion of an age-group in the population and additional life years left to calculate the weighted average of life expectancy by assigning weights according to the proportion in the population.

Supplementary Table 6: : Average life years saved

| **Age group**  **A** | **Deaths (%)**  **B** | **Average additional life years left.**  **C** | **Life years saved**  **D** |
| --- | --- | --- | --- |
| < 10 | 0.34 | 69.47 | 0.24 |
| 10-20 | 0.31 | 60.54 | 0.19 |
| 20-30 | 1.72 | 51.12 | 0.88 |
| 30-40 | 5.39 | 41.77 | 2.25 |
| 40—50 | 10.82 | 32.67 | 3.53 |
| 50—60 | 21.23 | 24.20 | 5.14 |
| 60—70 | 28.21 | 16.82 | 4.74 |
| 70—80 | 22.17 | 10.86 | 2.41 |
| > 80 | 9.81 | 6.150 | 0.60 |
| **Total life years saved** | | | 19.98 |

Notes: This table calculates life years saved by multiplying Deaths (%) and Average additional life years left and summing it up

for all age groups.

#### 1.4 Cost of estimation of public health interventions

**Cost of increased testing**

$$Cost incurred due to increased Tests, {IC}_{TT}=\left( \left( Total Tests \right)_{intervention}-\left( Total Tests \right)_{no intervention} \right)X C_{t}, where C_{t} is cost per RTPCR test$$

**Cost of Monitored Home Isolation**

The cost of monitoring home-isolated (MHI) patients is calculated by multiplying the total number of individuals in monitored home isolation by the daily per-patient monitoring cost. The total cost of monitoring isolated patients is determined by adding the daily cost to the doctor consultation cost over the entire epidemic duration. On average, patients with mild symptoms consult the doctor twice, while those with severe illness consult thrice during home-monitored isolation. Cost details are derived from discussions with a private institute collaborating with the government for monitoring home-isolated individuals.

$${Cost of Doctor Consultation, C}_{dc}=\left( \left( total mildly symptomatic patients in MHI X 2 \right) +\left( total severly ill patients in MHI X 3 \right) \right) X per Consultation charge$$

$$Cost ofMonitored Home Isolation(MHI),C_{mhi}=(\sum_{t=0}^{t_{end}} ({(total population in MHI)}_{t} X Cost of monitoring per patient per day))+C_{dc}$$

Cost of MHI per patient per day = Rs 46

Cost of per doctor consultation = Rs 100

**Cost of Facility based Isolation**

Using a calculator in [5], we obtained the inflation-adjusted cost in 2023 of critical care per patient per day, facility-based per patient per day in Kenya [6] and critical care per day per patient in India in 2021 [7] in US$. To account for the difference in standard of care in Kenya and India, we calculated cost of facility-based isolation by multiplying the ratio of cost of critical care per person per day in India to Kenya with cost of facility-based isolation per person per day in Kenya.

Patient cost-per day of critical COVID-19 care in Kenya, 2020= $ 599.51

Patient cost-per day of critical COVID-19 care in India, 2021= $ 210.99

Patient cost per day at isolation centre in Kenya, 2020 = $ 63.70

After, inflation adjusting all the costs to 2023, using inflation calculators [5]

cost of isolation centre per patient per day in India = $ 25.09

At exchange rate of 1US $ = 82.67 INR as of June 5, 2023, cost of isolation centre per patient per day is 2074.15 INR.

**Cost of hospitalisation**

Cost of hospitalization includes the include non-ICU admissions as well as ICU admissions accounted by severe and critical care patients respectively. Cost of severe care per person per day is calculated in the same manner as the cost of facility-based isolation per person per day. [8] estimates that 20.3% of hospitalised patients required ICU admissions. We calculate the average cost of hospitalisation per day by taking a weighted average of the cost of severe care and critical care per day according to their proportions.

$$Cost incurred due to increased hospitalization,{IC}_{H}=\left( \sum_{t=0}^{t_{end}} (\left( H_{1,0,t}+H_{1,2,t}+H_{2,0,t}+H_{2,1,t} \right)_{intervention} X hospitalization charges per day \right)- \left( \sum_{t=0}^{t_{end}} (\left( H_{1,0,t}+H_{1,2,t}+H_{2,0,t}+H_{2,1,t} \right)_{no intervention} X hospitalization charges per day \right)$$

With inflation adjusting and account for difference in standard of care in Kenya and India, we obtain that cost of severe-case care per day to be $ 236.81.

Cost of hospitalization per day = (0.797* $ 49.195) +(0.203*$ 236.81) = $ 87.28

At exchange rate of 1US $ = 82.67 INR as of June 5, 2023, cost of hospitalization per patient per day is 7215.437 INR.

### 2 Operational Model

#### 2.1 Sample Flow Calculation

**List of Abbreviations used in Operational Model.**

1. *N_cs_*_1_ = Number of Collection Sites per Sentinel Site (hub lab) = 6.
2. *N_cs_*_2_ = Number of Collection Sites per Sentinel Site (hospital) = 1.
3. *c_ss_*_1_ = Number of Sentinel Sites (hub lab) per Genome Sequencing Lab (GSL) = 5
4. *c_ss_*_2_ = Number of Sentinel Sites (hospitals) per GSL = 5.
5. *N_GSL_* = Number of GSLs = 57
6. *Total_cs_* = Total Number of collection sites = *N_GSL_* * (*c_ss_*_1_ * *N_cs_*_1_ + *c_ss_*_2_ * *N_cs_*_2_)
7. *N_ss_*_1_ = Set of Sentinel Sites (hub lab) = 1...... *N_GSL_* * *c_ss_*_1_ * *N_cs_*_1_ .
8. *N_ss_*_2_ = Set of Sentinel Sites (hospitals) = *(N_GSL_* * *c_ss_*_1_ * *N_cs_*_1_)+1.......*Total_cs_*
9. *Total_ss_* = Total Number of sentinel sites = *N_GSL_* * (*c_ss_*_1_ +*c_ss_*_2_ )
10. *t_cs_*_−_*_ss_* = Time taken (in days) to send samples from collection sites to sentinel sites = 1.
11. *t_SS_* = Time taken (in days) to extract RTPCR results = 1.
12. *Freq_SS_*_−_*_GSL_* = Frequency between samples sent from sentinel sites to GSL.
13. *t_SCAG_* = Time between two central advisory team meetings.
14. *t_PHR_* = Time taken (in days) by the state to start public health intervention since receiving the decision from NCDC = 15.
15. *Activities* = Number of activities performed in the sequencing lab.
16. *BS_i_* = Batch Size of Activity i.
17. *ProcessingTime_i_* = Processing Time of Activity i (mins/batch).
18. *I_vt_* = Number of samples of variant ’v’ collected on day t in the entire population across all collection sites. (**Input from EPI Model**)
19. *I*‘_2_*_t_* = Number of samples of *I*_2_*_t_* entering into WGS network. **(Random variable drawn from hypergeometric distribution)**
20. *SCT_vcjkt_* = Number of samples of variant v collected for testing at collection site c under sentinel site j, GSL k at time t.
21. *SRT_vjkt_* = Number of samples of variant v received for testing at sentinel site j under GSL k at time t.
22. *T_vjkt_* = Number of samples of varinat v tested at sentinel site j under GSL k at time t.
23. *P_ct_* = Proportion of positive samples with ct value ≤ 25. = 0.75
24. *EGSL_vjkt_* = Number of samples of variant v eligible to be sent to GSL k from sentinel site j at time t.
25. *SGSL_jkt_* = Total number of samples sent to GSL k from sentinel site j at time t.
26. *SGSL*_1_*_jkt_* = Number of samples of variant 1 sent to GSL k from sentinel site j at time t.
27. *SGSL*_2_*_jkt_* = Number of samples of variant 2 sent to GSL k from sentinel site j at time t. **(Random variable drawn from hypergeometric distribution)**
28. *NS_SS_*_−_*_GSL_* = number of samples to be sent from sentinel site to GSL.

(5%, 10%,30% ,… of eligible samples)

29. *SRS_vkt_* = Number of samples of variant v received for sequencing at GSL k at time t.
30. *S_kt_* = Number of samples sequenced at GSL k at time t.
31. *Inv_ikt_* = Number of samples in the inventory before each activity i at GSL k at time t.
32. *Ex_ikt_* = Number of samples exiting from each activity i at GSL k at time t.
33. *MI_ikt_* = Number of machines idle before each activity i at GSL k at time t.
34. *MU_ikt_* = Number of machines in use before each activiy i at GSL k at time t.

**Number of samples of variant 2 entering the WGS network on each day.**

The number of samples of variant 2 entering the WGS network on each day to be a random variable drawn from a hypergeometric distribution with probability,

P(X = *x*) = $\frac{a_{C_{x}} * {(n- a)}_{C_{(r- x)}}}{n_{C_{r}}}$

Where, x = *I*‘_2_*_t_* = number of samples of variant 2 entering WGS network

a = *I2t*

n = *I1t + I2t*

r = n * 8% = Total number of samples entering into the WGS network.

The number of samples of variant 1 entering the WGS network on each day will be estimated by subtracting the number of samples of variant 2 that enter the network each day from the total number of samples entering into the WGS network each day.

*I*‘_1_*_t_*	= r - *I*‘_2_*_t_*

The variant 1 and variant 2 samples collected could be part of any of the 1995 collection sites associated under 57 decentralized GSL or associated under 1 Centralized GSL. There will be ${(n+k- 1)}_{C_{(n- 1)}}$ possible combinations of ‘n’ samples (variant 1 or variant 2) assigned to k collection sites. We choose 1 of these possibilities for each variant to track which collection site the sample of variant 1 and variant 2 are part of.

**Number of samples tested each day**

The number of samples of variant v received at a sentinel site (hub lab) each day is estimated by the

sum of samples collected for testing across all collection sites under the hub sentinel lab.

SRT*_vjkt_* = $\sum_{c\in Ncs1} SCTvct$ ∀*v* ∈ (1*,*2) *j* ∈ *Nss*1*, k* ∈ *NGSL, t > tcs*−*ss*

The number of samples of variant v received at a sentinel site (hospital) each day is estimated by the

sum of samples collected for testing across all collection sites within the hospital.

SRT*_vjkt_* = $\sum_{c\in Ncs2} SCTvct$ ∀*v* ∈ (1*,*2) *j* ∈ *Nss*2*, k* ∈ *NGSL, t > tcs*−*ss*

Initializing the SRT*_vjkt_* and T*_vjkt_*

SRT*_vjkt_* = 0	∀*v* ∈ (1*,*2) *j* ∈ *Total_ss_	k* ∈ *N_GSL_,t* ≤ *t_cs_*_−_*_ss_*

T*_vjkt_* = 0	∀*v* ∈ (1*,*2) *j* ∈ *Totalss	k* ∈ *NGSL,t* ≤ *tcs*−*ss* + *tss*

The number of samples tested at a sentinel site each day are the samples that were received *tcs*−*ss* + *tss* days

ago.

T*_vjkt_* = SRT*_vjkt_* *_−(tcs−ss+tss)_* ∀*v* ∈ (1*,*2)*, j* ∈ *Totalss, k* ∈ *NGSL,t > tcs*−*ss* + *tss*

**Number of samples i) eligible to be sent to GSLs and ii) sent to GSLs**

The total number of samples sent to GSL from a sentinel site during each shipment is estimated by the minimum of eligible samples at a sentinel site and proportion of eligible samples that needs to be sent to GSL.

$${SGSL}_{jkt}= \left\{ \begin{aligned} \min\left( \sum_{v \in\left( 1,2 \right)} {EGSL}_{vjkt},{NS}_{SS-GSL}* {EGSL}_{vjkt} \right), &t=a *\left( \frac{1}{{Freq}_{SS-GSL}} \right), a=1,2,3\ldots.n \\ \\ \\ \\ 0, otherwise \end{aligned} \right.$$

The number of samples that are eligible to be sent to GSL each day from hub sentinel labs

EGSL*_vjkt_* = *T_vjkt_* ∗ *Pct* + *EGSL_jkt−1_* − *SGSL_jkt−1_*	∀*v* ∈ (1*,*2) *,j* ∈ *N_ss_*_1_*, k* ∈ *N_GSL_	,t >* 0

The number of samples that are eligible to be sent to GSL each day from hospitals

EGSL*_vjkt_* = *T_vjkt_* ∗ *Pct* + *EGSL_jkt−1_* − *SGSL_jkt−1_*	∀*v* ∈ (1*,*2) *,j* ∈ *N_ss_*_2_*, k* ∈ *N_GSL_	,t >* 0

**Number of samples of variant 2 sent to an GSL from each sentinel site.**

The number of samples of variant 2 sent to an GSL is a random variable drawn from a hypergeometric distribution with probability,

P(X = x`) = $\frac{{a`}_{C_{x`}} * {(n`- a`)}_{C_{(r`- x`)}}}{{n`}_{C_{r`}}}$

Where, x` = SGSL*_2jkt =_* number of samples of variant 2 sent to GSL during each shipment.

a` = EGSL*_2jkt_*

n` = EGSL*_1jkt_* + EGSL*_2jkt_*

r` = min (5% or 10% or 30%, n) = Total number of samples sent to GSL during each shipment.

On each shipment day which is 7 days the number of samples of variant 1 sent to an GSL will be estimated by subtracting the number of samples of variant 2 that are sent to the GSL from the total number of samples that are sent to the GSL during each shipment.

SGSL*_1jkt =_* r` - SGSL*_2jkt_*

**Number of samples sequenced at an GSL**

The number of samples received for sequencing at an GSL from all sentinel sites under it.

SRS*_vkt_* = $\sum_{j\in Nss1} {SGSL}_{vjkt}$+ $\sum_{j\in Nss2} {SGSL}_{vjkt}$ ∀*v* ∈ (1*,*2) *k* ∈ *N_GSL_ , t >* 0

For each of the activities we first capture the inventory of samples left or available to be processed each day. This is done by adding the number of samples processed (exiting the activity) in the previous activity on a given day and the number of samples in the inventory to be processed in the current activity the previous day then subtracting the number of samples processed by the current activity the previous day.

The Inventory for the first activity at an GSL each day

*Inv*1*,k,t* = $\sum_{v\in(1,2)} SRSvkt$ + *Inv*1*,k,t*−1 − *Ex*1*,k,t*−1	∀*k* ∈ *NGSL	t >* 0

The number of samples in inventory for each activity (2nd activity onwards) at an GSL each day.

*Invi,k,t* = *Exi*−1*,k,t* + *Invi,k,t*−1 − *Exi,k,t*−1 ∀*i* ∈ *Activities , k* ∈ *NGSL, t >* 0

The number of machines idle for an activity at an GSL each day.

*MIi,k,t* = Total number of machines available for the activity - *MUi,k,t* ∀*i* ∈ *Activities , k* ∈ *NGSL, t >* 0

The number of machines in use for an activity at an GSL each day. This checks how many of machines are currently in use for an activity.

*MUi,k,t* = $\left( \frac{{BP}_{ik (t-{ProcessingTime}_{i})\ldots(t-1)}}{{BS}_{ik}} \right)$ ∀*i* ∈ *Activities , k* ∈ *NGSL, t >* 0

Samples processed from an acitvity on each day.

*BPi,k,t* = min $\left( {MI}_{ikt} * {BS}_{ik} , \left( \frac{Invi,k,t}{{BS}_{ik}} \right)* BSi,k \right)$ ∀*i* ∈ *Activities , k* ∈ *NGSL, t >* 0

On a given day if samples are processed they exit the activity after the processing time. The number of samples exiting an activity at an GSL each day,

${Ex}_{ikt}= {BP}_{ik (t-{ProcessingTime}_{i})}$	∀*i* ∈ *Activities , k* ∈ *NGSL, t >* 0

The number of samples sequenced each day at an GSL is the number of samples exiting the last activity

each day.

*S_kt_* = *Ex_ikt_*	∀*i* ∈ *last Activity in the lab	k* ∈ *N_GSL_*

In the model, we keep track of the entry time of the first sample of variant 2 in the network (t1) i.e., the time when the sample of variant 2 is collected for testing at a collection site part of the WGS network and also the exit time of the first sample of variant 2 in the network (t2). i.e., the time when the sample is sequenced and uploaded to Genome Sequence Database (GSD).

For each simulation, the turnaround time of implementing an intervention is estimated by the difference between t1 and variant emergence time (Vet) is added to the difference between t2 and t1+ the average time taken to share the sequenced results with the central advisory team+ Time taken by the state to implement the intervention .

TAT = (Vet – t1) + (t2 – t1) + $\left( \frac{t_{SCAG}}{2} \right)$ + $t_{PHR}$

We run the model for 100 simulations and estimate the average turnaround time of implementing an intervention.

#### 2.2 Cost estimation of Operation

**Cost of Shipment:**

The total cost of shipment is the summation of the cost of shipment incurred each week to send samples from sentinel sites to GSL. The cost of shipment included variable expenses for packing materials (Viral Transport Medium - VTMs, secondary container pouches, insulation boxes) dependent on the sample quantity per shipment, along with a fixed transportation cost (courier charges) for delivering samples to GSL. Each sample required a VTM and secondary container pouch. The count of insulation boxes was calculated by dividing the total packed storage volume of all samples by the storage volume of each box. Our investigation revealed a fixed transportation cost for every 100 samples. Shipment cost was determined by rounding up the samples for shipment, dividing by 100, and multiplying by the fixed transportation cost per 100 samples. We assume that in a decentralized network, road services were used, while air services transported samples in a centralized setup. To estimate air transportation cost, we utilized the air-to-road cost ratio from BlueDart courier service, which was based on a fixed origin-destination pair for sending a volume equivalent to 100 samples. For a centralized network, transportation cost was approximated by multiplying the road transportation cost for 100 samples by this ratio (see **Supplementary Table 8** and **Supplementary Table 8**).

Supplementary Table 7: Shipment Cost Calculations and Estimates

| **Average # of samples**  **sent per shipment**  **A** | **Transportation Cost (INR)**  **B = (A/100)*1000^b^** | **Packaging Cost (INR)**  **C = (A/40)*250^c^** | **Material Cost (INR)**  **D = A*60^d^** | **Average Cost per Shipment (INR)**  **(B+C+D)** |
| --- | --- | --- | --- | --- |
| 20 | 1000 | 250 | 1200 | 2450 |
| 50 | 1000 | 500 | 3000 | 4500 |
| 100 | 1000 | 750 | 6000 | 7750 |
| 150 | 2000 | 1000 | 9000 | 12000 |
| 200 | 2000 | 1250 | 12000 | 15250 |

Notes:

a. Costs incurred in transportation, packaging and materials were obtained from interviews with officials who were part of sending samples to sequencing labs.
b. INR 1000 is inccured to send upto 100 samples from source to destination under decentralized network, whereas INR 3168 is incurred under centralized network.
c. Cost of one thermocol box and dry ice placed in each thermocol box. Also, each thermocol box can hold upto 40 samples.
d. Cost of one VTM and one secondary container**.**

Supplementary Table 8: Additional Costing parameters

|  |  | **Values** | **Source** |
| --- | --- | --- | --- |
| A1 | Volume per sample (in ml) | 2 | Noted from Interview with official involved in WGS implementation |
| A2 | Number of VTM's required per sample | 1 | [9] |
| A3 | Volume of each secondary storage container (in ml) | 50 | [9] |
| A4 | Number of samples stored in secondary storage container | 1 | [9] |
| A5 | Packed storage volume per sample (in ml) | 50 | A3/A4 |
| A6 | Gross Volume of thermocol box (ml) | 3000 | Noted from Interview with official involved in WGS implementation |
| A7 | Net Volume of thermocol box (ml) | 2010 | A6 * 67% utilization factor [10] |
| A8 | Number of samples placed in each thermocol box | 40 | A7/A5 |
| A9 | Number of thermocol boxes required to ship 100 samples | 3 | 100/A8 |
| A10 | Total volume of shipment (in ml) | 9000 | A9 * A6 |
| A11 | Cost of Transportation (courier charges) of 100 samples by road - decentralized network (INR) | 1000 | Noted from Interview with official involved in sending samples to GSL |
| A12 | BlueDart courier charges to ship a volume of 9L by road (INR) | 970 | https://www.bluedart.com/home#transitfinder |
| A13 | BlueDart courier charges to ship a volume of 9L by air (INR) | 3073 | https://www.bluedart.com/home#transitfinder |
| A14 | Cost of Transportation (courier charges) of 100 samples by road - decentralized network (INR) | 3168 | A11 * (A13/A12) |
| A15 | Cost per thermocol box (INR) | 50 | Noted from Interview with official involved in WGS implementation |
| A16 | Cost of Dry Ice (INR) | 200 | Noted from Interview with official involved in sending samples to GSL |
| A17 | Cost of each VTM (in Rs.) | 30 | https://www.indiamart.com/proddetail/abi-viral-transport-medium-vtm-kit-22559755048.html?pos=16 |
| A18 | Cost of each secondary container (in Rs.) | 30 |  |
| A19 | Material Cost per Sample | 60 | A17 * A2 + A18 * A4 |
| A20 | Packaging Cost per thermocol box | 250 | A15 + A16 |

Notes: Values marked in orange are used in the calculations of **Supplementary Table 7**

**Cost of Personnel and Reagents:**

Activity-based costing was used to estimate the costs of personnel and reagents used at an GSL. The time taken by personnel per batch for each activity was noted during our field visit to an GSL. The number of reagents used either per sample or per batch in each activity and salary of personnel is obtained from interviews while [11] listed the cost of Illumina reagent kits.

The total cost of personnel and reagents was estimated by multiplying the total number of runs required to sequence all the required samples. The number of reagent kits required per activity is estimated and multiplied by the cost of each reagent kit. The cost of personnel incurred per sequencing run is estimated by considering the time taken (in hours) by personnel in each activity and multiplied by the salary given to the personnel for a batch of 384 samples. (Refer **Supplementary Table 9** **and Supplementary Table 10** for details)

Supplementary Table 9 Activity-wise reagents used and respective unit costs.

| **Activity** | **Reagent Used** | **Number of Samples processed per Reagent Kit** | **Unit Price per Reagent Kit (INR)^c^** | **Cost incurred per sequencing run** |
| --- | --- | --- | --- | --- |
| Sequencing^ab^ | MiSeq Reagent and Flow Cell | 384 | 132819.5 | 132819.5 |
| RNA to cDNA | AmpliSeq cDNA Synthesis | 100 | 41738 | 160273.9 |
| Library Prep | Illumina DNA Prep | 96 | 323518.7 | 1294075 |
| Library Prep | Illumina DNA Prep Kit Indexes | 96 | 38482.6 | 153930.4 |

Notes

a. Reagent Kits for Sequencing Activity are used per run basis whereas for all other activities they are used per sample basis.
b. Sequencer used : MiSeq
c. Unit price per reagent kit (INR) are taken from Illumina’s quotation to the state of Maine, United States of America [11].

Supplementary Table 10 Activity wise time taken by personnel and respective unit costs

| **Activity** | **Time Taken to process**  **one batch (in mins)** | **Batch Size of Personnel per activity** | **Personnel Involved** | **Incentive per hour**  **(INR)** | **Cost of Personnel per Activity (INR)** |
| --- | --- | --- | --- | --- | --- |
| Sample Reception | 1 | 1 | Lab Associate | 175 | 1120 |
| RNA to cDNA | 1 | 1 | Lab Associate | 175 | 1120 |
| Library Prep | 1.85 | 1 | Lab Associate | 175 | 2072 |
| Sequencing | 1 | 1 | Lab Scientist | 450 | 2880 |
| Bioinformatics | 0.62 | 1 | Lab Scientist | 450 | 1785.6 |

Notes:

Cost of personnel per activity

= (# of samples to be sequenced per run / Batch Size of personnel per activity) * Time taken to process one batch (in Hrs.) x Salary per hour

Sample Reception Example  = (384/1) * (1/60) * 175 =  Rs. 1120

**Cost of Equipment:**

The list and purchase cost of each equipment item were collected from interviews. The annual cost of each equipment was calculated by dividing the purchase cost of each equipment by an annualization factor [12]. The annualization factor is estimated by considering a discount rate of 3.5% and a life span of 5 years per equipment [13] (Refer **Supplementary Table 11** for details). The equipment costs were then weighted based on the percentage of time that each equipment item was used for genome sequencing. It was assumed that all the equipment items are exclusively used for genome sequencing, with 100% of their time dedicated to WGS. The cost of equipment per week was estimated by dividing the annual cost by the number of working weeks in a year. The total cost of equipment is estimated by multiplying the equipment cost per week by the total number of weeks taken to complete sequencing. (Refer **Supplementary Table 9** for details)

Supplementary Table 11: Activity wise Equipment’s used and respective unit costs

| **Actvity** | **Equipment Used** | **Initial Purchase Cost (INR)** | **Annual Cost^c^ (INR)** | **Number of Equipments in the lab** | **Cost of the Equipment per week** |
| --- | --- | --- | --- | --- | --- |
|  |  | A | B = A / A.F | B | (A*B)/number of working weeks in a year |
| Sequencing | MiSeq* | 4500000 | 996666.2 | 1 | 23178.3 |
| RNA to cDNA | 96 well PCR system | 260000 | 57585.2 | 3 | 4017.6 |
| Library Prep | Storage server: 16 GB RAM, 8 Core Processor,4 TB Storage | 200000 | 44296.3 | 1 | 1030.1 |
| Library Prep | UPS (2 KVA) 30 Min back up | 100000 | 22148.1 | 1 | 515.1 |
| RNA to cDNA | Microplate centrifuge, PCR plate spinner* | 150000 | 33222.2 | 1 | 772.6 |
| RNA to cDNA | MiniSpin(Quick Spin 1.5 ml MCTs) | 15000 | 3322.2 | 1 | 77.3 |
| RNA to cDNA | Mini Spin (Quick Spin 8 tube strips) | 15000 | 3322.2 | 1 | 77.3 |
| Library Prep | Magnetic stand-96 well plate | 45000 | 9966.7 | 2 | 463.6 |
| Library Prep | Magnetic stand 1.5/2 ml tubes | 55000 | 12181.5 | 1 | 283.3 |
| Library Prep | Vortex | 18000 | 3986.7 | 2 | 185.4 |
| Library Prep | Autopipette set to handle 0.5 µL to 1 mL | 30000 | 6644.4 | 2 | 309.0 |
| Library Prep | 8- channel autopipette 30 - 300 µL and 0.5- 10 µL (one each) | 52620 | 11654.3 | 2 | 542.1 |
| Library Prep | Qubit Fluorometer* | 300000 | 66444.4 | 1 | 1545.2 |
| Library Prep | Bio Shake system with plate adapter | 400000 | 88592.5 | 1 | 2060.3 |
| RNA to cDNA | PCR hood | 45000 | 9966.7 | 1 | 231.8 |
| Library Prep | Vibration Free table | 40000 | 8859.3 | 1 | 206.0 |
| BioInformatics | Initial Software Cost | 250000 | 50000.0 | 1 | 1162.8 |
| BioInformatics | Software Cost Subscription | 50000 | 50000.0 | 1 | 1162.8 |

Notes:

a. Number of working weeks in a year is considered to be 43 weeks.
b. List of equipment used in a sequencing lab and their initial purchase cost were obtained from interviews with officials who were part of establishing WGS labs.
c. Annual Cost (INR) of each equipment is estimated by divinding the initital purchase cost of the equipment (INR) by an annualization factor (A.F). A.F is given by [(1+r)^n^ – 1]/[r(1+r)^n^] where r is the discount rate and n is the life years of the equipment [12]. For a discount rate (r) of 3.5% and 5 life years, the annualization factor is 4.51505

The number of Sequencing, Centrifuge and Qubit machines increase on increasing capacity. In our model, increasing sequencing capacity by 5 times, requires 4 additional sequencing machines, 2 additional centrifuge and qubit machines. Hence, an additional INR 97,348.8 will be incurred per week when the capacity is increased

**Cost of RTPCR Tests:**

The total cost of RTPCR tests is estimated by multiplying the number of samples that are tested again at an GSL with the cost of an RTPCR test. We assume that 5% of samples that reach GSL each week are retested. [14].

### 3 Supplementary Figures

Supplementary Figure 1: Exposed and infectious compartments incorporating RT-PCR test status

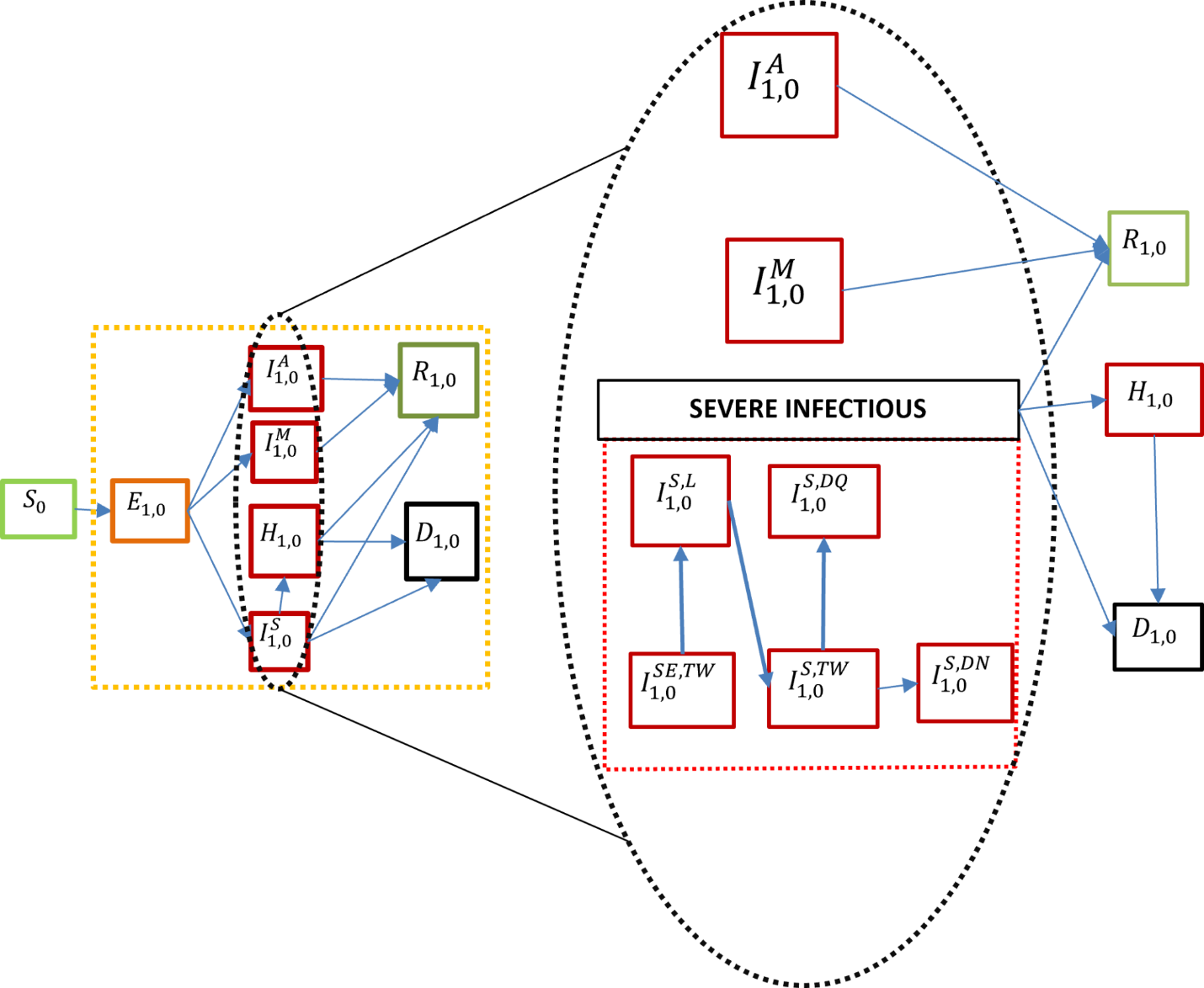

Note: This figure represents different compartments of epidemiological model and Severely-ill Infectious disease state being divide into multiple compartments based on RTPCR results status. Refer to supplementary section 1.1 for detailed description of each compartment.

Supplementary Figure 2: Daily infections from calibrated model and OWID dataset

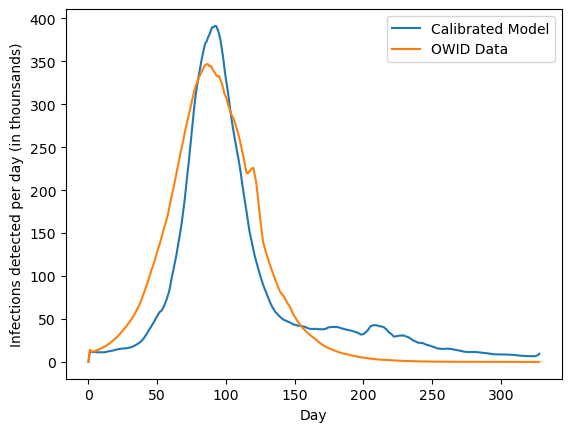

Note:
This figure represents the calibration of the epidemiological model using observed case data. The x-axis shows days since the start of the simulation, and the y-axis shows the number of infections detected per day (in thousands). The blue line represents infections predicted by the calibrated model, while the orange line represents reported daily infections from the Our World in Data (OWID) dataset between February 2021 and December 2021.

Supplementary Figure 3: Model vs. Data Average CST comparison

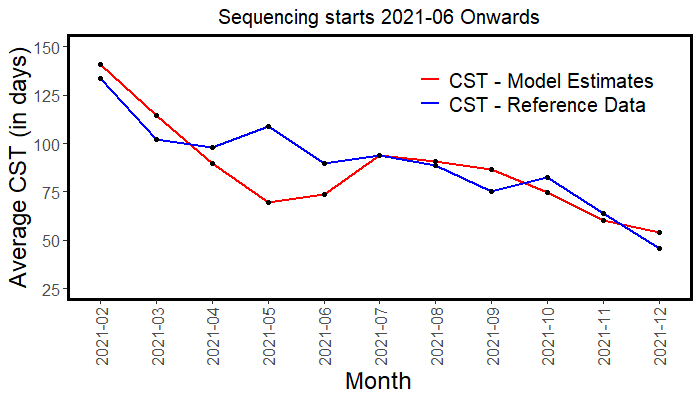

Supplementary Figure 4: Varying sequencing capacity estimated from the model

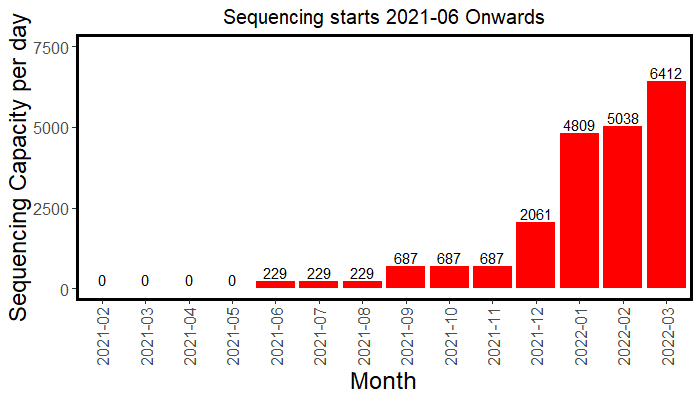

Supplementary Figure 5: Infections averted in scenarios at different operatinal cnfigurations
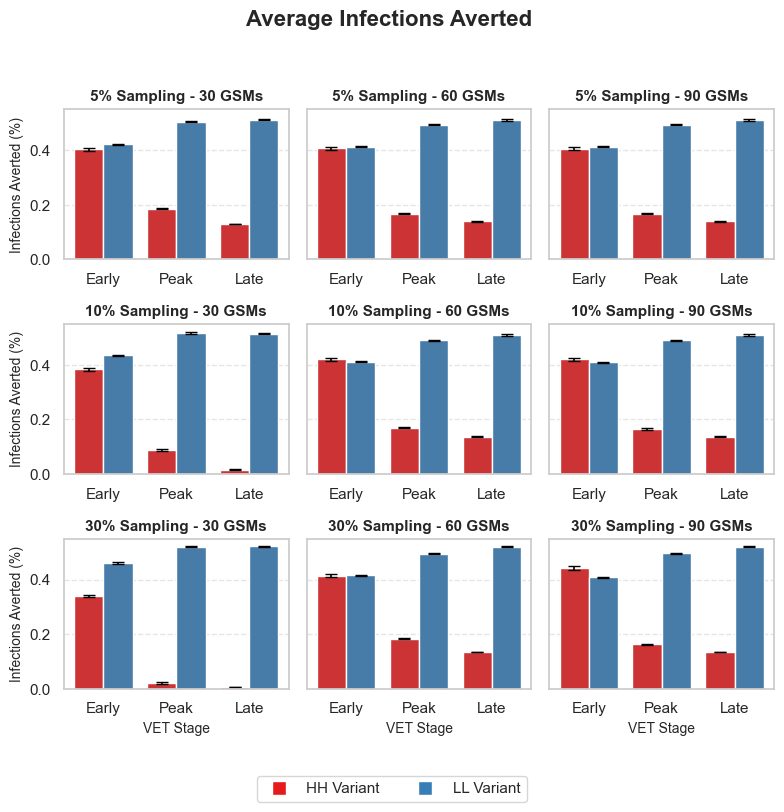

Note:
This figure represents the average percentage of infections averted due to intervention across different operational and epidemiological scenarios. The x-axis denotes the variant emergence timepoint (VET Stage: Early, Peak, Late), and the y-axis shows the percentage of infections averted. Each subplot column corresponds to a fixed number of Genome Sequencing Machines (GSMs): 30 GSMs (first column), 60 GSMs (second column), and 90 GSMs (third column). Each subplot row corresponds to a sampling proportion of positive cases for genome sequencing: 5% (first row), 10% (second row), and 30% (third row). Red bars represent the High severity–high immune escape (HH) variant, and blue bars represent the Low severity–low immune escape (LL) variant.

Supplementary Figure 6: Deaths averted in differetn scenarios at different operational configurations

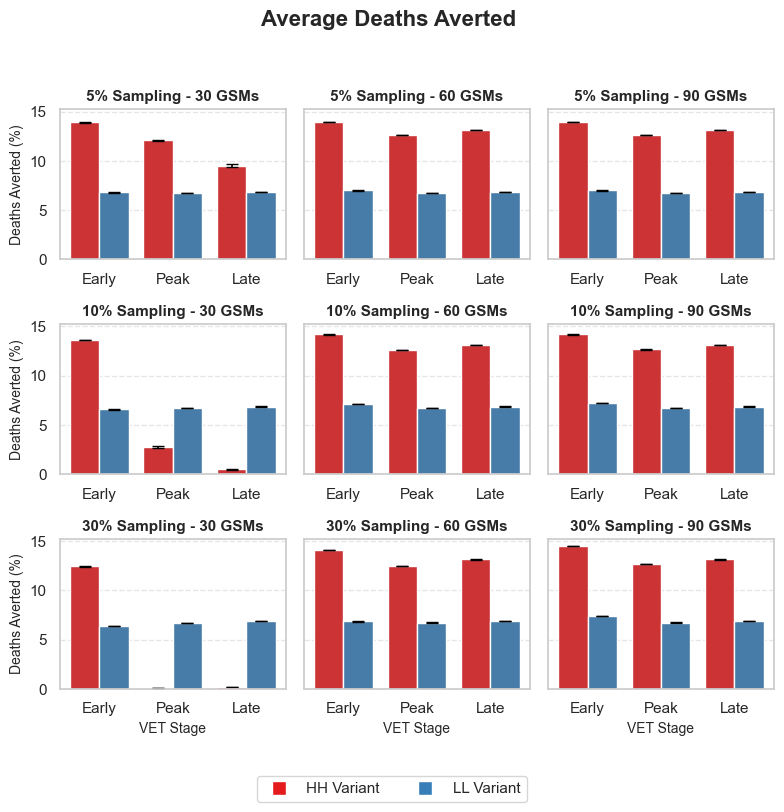

Note:
This figure represents the average percentage of deaths averted due to intervention across different operational and epidemiological scenarios. The x-axis denotes the variant emergence timepoint (VET Stage: Early, Peak, Late), and the y-axis shows the percentage of deaths averted. Each subplot column corresponds to a fixed number of Genome Sequencing Machines (GSMs): 30 GSMs (first column), 60 GSMs (second column), and 90 GSMs (third column). Each subplot row corresponds to a sampling proportion of positive cases for genome sequencing: 5% (first row), 10% (second row), and 30% (third row). Red bars represent the High severity–high immune escape (HH) variant, and blue bars represent the Low severity–low immune escape (LL) variant.

### 4 Supplementary Tables

Supplementary Table 12: List of activities in the sequencing lab with batch size and processing time

| **Sl No** | **Activity** | **Batch Size^a^** | **Processing Time per batch**  **(in mins)** |
| --- | --- | --- | --- |
| 1 | Aliquoting | 1 | 1 |
| 2 | RNA to cDNA conversion | 96 | 60 |
| 3 | Barcoding | 1 | 1 |
| 4 | Multiplexing | 1 | 0.63 |
| 5 | Quality Check | 96 | 60 |
| 6 | Pooling | 1 | 0.47 |
| 7 | Sequencing | 384 | 720 |

Notes

a. The batch size for all activities indicates the maximum batch size that can be processed in one go.

Supplementary Table 13: Parameters of WGS Surveillance Network associated with one GSL

| **Sl No** | **Parameter** | **Value^a^** |
| --- | --- | --- |
| 1 | Number of Collection Sites per Sentinel Site (hub lab) | 6 |
| 2 | Number of Collection Sites per Sentinel Site (hospital) | 1 |
| 3 | Number of Sentinel Sites (hub lab) per GSL | 5 |
| 4 | Number of Sentinel Sites (hospitals) per GSL | 5 |
| 5 | Number of GSLs^b^ | 57 |
| 6 | Transportation frequency between collection sites and sentinel sites | every day |
| 7 | Transportation frequency between sentinel sites and GSL | every 7 days |
| 8 | Frequency of central advisory team meetings | every 7 days |
| 9 | Number of samples sent from sentinel site to GSL | eligible samples per sentinel site * Sample Proportion |

Notes

a. Values of parameters 6 – 9 are subject to operational decisions. They may be varied as per the situational demand. In our model, we vary the number of samples sent from the sentinel site to GSL ( 5%, 10%, 30% of eligible samples )
b. Values of the number of GSLs are specific to the decentralized sequencing network scenario.

[9]	Sumit. Title: Specimen Collection, Packaging and Transport Guidelines for 2019 Novel Coronavirus (2019-nCoV). n.d.

[10]	WHO Vaccine Management Handbook Module VMH-E3-01.1 HOW TO CALCULATE VACCINE VOLUMES AND COLD CHAIN CAPACITY REQUIREMENTS EVM Setting a standard for the vaccine supply chain. 2017.

[11]	Contract Web Doc n.d.

[12]	Walker D, Kumaranayake L. Introduction: costs over time How to do (or not to do). .. Allowing for differential timing in cost analyses: discounting and annualization. 2002.

[13]	Schwarze K, Buchanan J, Fermont JM, Dreau H, Tilley MW, Taylor JM, et al. The complete costs of genome sequencing: a microcosting study in cancer and rare diseases from a single center in the United Kingdom n.d. https://doi.org/10.1038/s41436.

[14]	Minhas N, Gurav YK, Sambhare S, Potdar V, Choudhary ML, Bhardwaj SD, et al. Cost-analysis of real time RT-PCR test performed for COVID-19 diagnosis at India’s national reference laboratory during the early stages of pandemic mitigation. PLoS One 2023;18. https://doi.org/10.1371/journal.pone.0277867.
